# Reperfusion strategies in patients with ST-segment elevation myocardial infarction during hospitalization in China: Findings from the Improving Care for Cardiovascular disease in China (CCC)-Acute Cronary Synrome project

**DOI:** 10.1101/2023.12.10.23299554

**Authors:** Jun Wang, Zhiqiang Zhang, Jing Li, Xiaoxiang Tian, Xiaozeng Wang, Yaling Han, CCC investigators

## Abstract

**Objective:** To analyze the current situation of reperfusion strategies of ST-segment elevation myocardial infarction (STEMI) in China and evaluate the efficacy and safety of different reperfusion strategies, especially pharmaco-invasive percutaneous coronary intervention (PI-PCI).

**Methods:** The CCC-ACS (Improving Care for Cardiovascular Disease in China-Acute Coronary Syndrome) project is a joint study between the American Heart Association and Chinese Society of Cardiology (CSC). STEMI patients who were recruited to the CCC-ACS project between November 2014 and December 2019 and admitted within 48 hours after symptom onset and treated by thrombolysis or percutaneous coronary intervention (PCI) were included in this cohort study. The primary efficacy outcomes were major adverse cardiac cerebrovascular events (MACCEs) that occurred during hospitalization. The primary safety outcomes were Thrombolysis in Myocardial Infarction (TIMI) major or minor bleedings criteria during hospitalization. Univariate regression logistic analysis, multivariable logistic regression analysis, propensity score-matched analysis, and inverse probability of treatment weighting analysis were performed to evaluate the efficacy and safety of different reperfusion strategies.

**Results:** Of 37733 STEMI patients, 35019 patients received primary percutaneous coronary intervention (PPCI), 999 patients received thrombolysis and 1715 patients received PI-PCI. Compared with PPCI, the thrombolysis group had higher incidence of all cause death (1.6% vs 2.8%, P =0.003), MACCEs (2.0% vs 3.6%, P < 0.001), and TIMI major bleedings (1.2% vs 2.2%, P=0.007). In the PI-PCI group, the incidence of MACCEs (2.0% vs 0.8%, P =0.001), all cause death (1.6% vs 0.4%, P =0.001), and cardiac death (1.5% vs 0.4%, P =0.001) were significantly lower than PPCI group; and the same conclusion was found in the subgroup of in time from first medical contact(FMC) to reperfusion ≥ 3h. However, the risk of TIMI minor bleedings (5.1% vs 6.7%, P=0.008) was higher in the PI-PCI group in the subgroup of in time from FMC to reperfusion ≥ 3h. Compared with timely PPCI group, the incidence of all cause death was significantly lower and the incidence of heart failure was higher in the scheduled PCI group. Compared with late PPCI group, the incidence of all cause death, MACCEs were significantly lower in scheduled PCI group. Compared with timely PPCI, the ratio of heart failure was statistically significant higher in the rescue PCI group. There was no significant difference in all outcomes in all models between rescue PCI group and late PPCI group. Moreover, compared with scheduled PCI ≤ 24h group, the scheduled PCI during 24h to 7d group had lower risk of TIMI major or minor bleedings and the scheduled PCI >7d group had the similar risk of bleedings; the scheduled PCI >7d group had lower risk of heart failure.

**Conclusions:** This study demonstrates that in STEMI patients who could not perform timely PPCI, PI-PCI is feasible, including rescue PCI,which can reduce the rate of MACCEs and mortality during hospitalization.But the increased risk of bleedings also should be noted.In scheduled PCI after successful thrombolysis, appropriate extension the time window of scheduled PCI can be considered under stable clinical conditions.

## 1. Introduction

ST-segment elevation myocardial infarction (STEMI) is a clinical syndrome defined by the presence of myocardial ischemic symptoms, electrocardiographic (ECG) findings of new ST-segment elevations in two continuous leads or new left bundle branch block, and subsequent detection of biomarkers indicative of myocardial injury.^1^

China is facing the dual pressure of aging population and continuous prevalence of metabolic risk factors, and the burden of cardiovascular diseases and the mortality of acute myocardial infarction is increasing.^2^ Primary percutaneous coronary intervention (PPCI) has been the preferred reperfusion strategy for patients with STEMI. However, PPCI is not universally available, and delays in performing PPCI are common in real-world practice.^3^ Even in some large cities, patients have a high chance of presenting to hospitals not providing around-the-clock PPCI service. The outcome of STEMI is varied significantly in different hospitals. As the efficacy of PPCI is time-dependent, the no-reflow phenomenon in PPCI may lead to failure of myocardial reperfusion. Therefore, intravenous thrombolysis still remains a viable option for reperfusion, and plays an important role in modern STEMI management. However, the reoccurrence of myocardial ischemia after thrombolytic therapy is common. Pharmaco-invasive percutaneous coronary intervention (PI-PCI) strategy, an early reperfusion strategy encompassing initial prompt fibrinolysis with subsequent early catheterization, has been proposed as a therapeutic option for STEMI patients when timely PPCI is not available.^4,5^

Current evidence on the efficacy and safety of the therapeutic strategies for PI-PCI in patients with STEMI remains limited. We aim to analyze the current situation of reperfusion strategies of STEMI in China and compare the efficacy and safety of different reperfusion strategies during hospitalization, especially PI-PCI.

## 2. Methods

### 2.1 Study design and patient selection

The CCC-ACS (Improving Care for Cardiovascular Disease in China-Acute Coronary Syndrome) project, a nationwide registry and quality improvement study focusing on quality of acute coronary syndrome (ACS) care, was launched in 2014 as a collaborative initiative of the American Heart Association and the Chinese Society of Cardiology across China. This study is registered at ClinicalTrials.gov (unique identifier: NCT02306616) and complies with the Declaration of Helsinki. Detailed information on the design and methodology of the CCC-ACS project has been published previously.^6^ STEMI was defined in accordance with the Chinese Society of Cardiology guidelines for the diagnosis and management of patients with non-ST-segment elevation ACS and STEMI.^7, 8^ From November 1, 2014 to December 30, 2019, a total of 113,651 patients with ACS were enrolled in the CCC-ACS project. Among them, 37733 inpatients with STEMI admitted within 48 hours after symptom onset and treated by thrombolysis or PCI were selected for analysis. The inclusion and exclusion criteria are shown in Figure 1.

**Figure 1.**
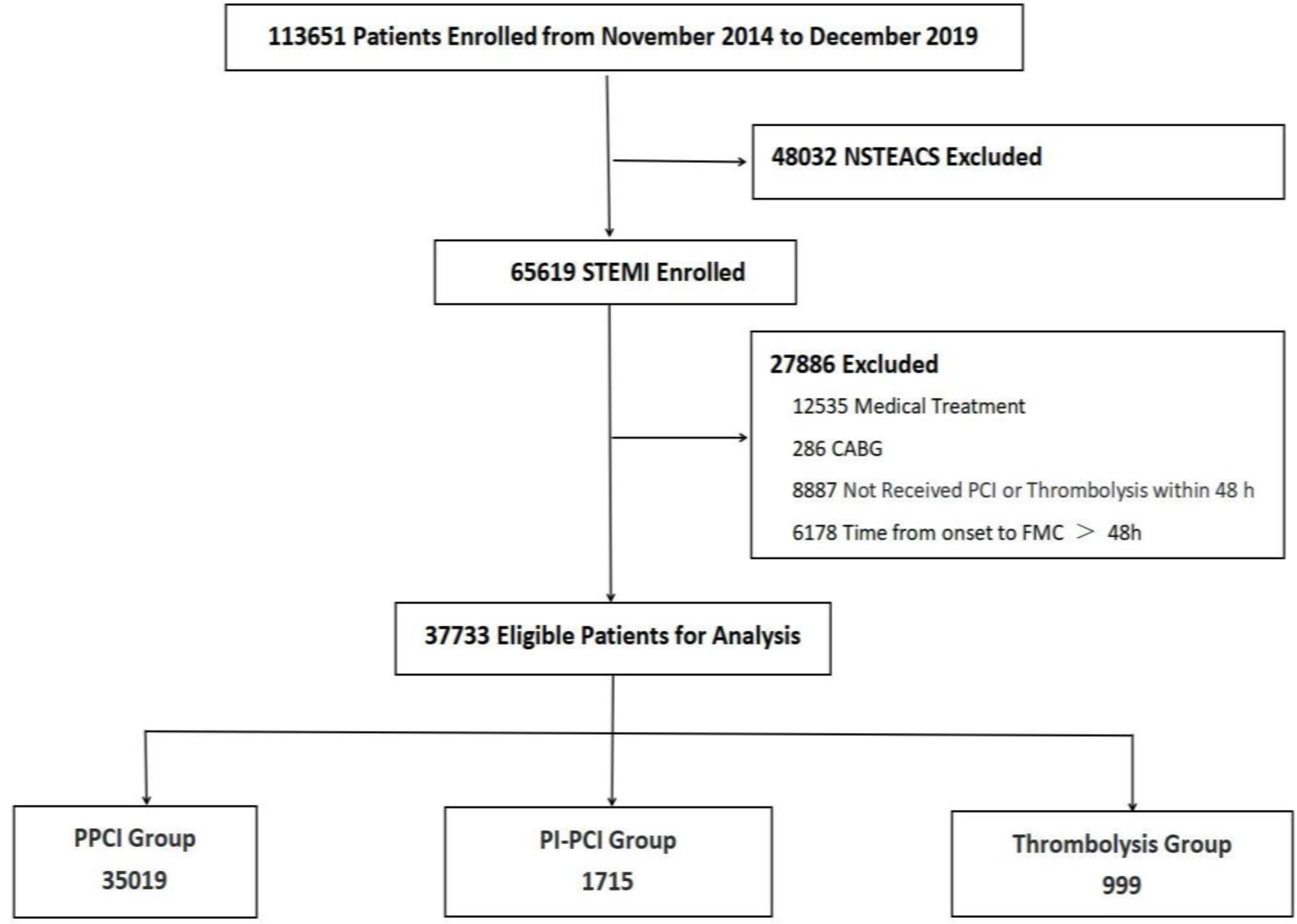
Study flowchart PCI=percutaneous coronary intervention; PPCI=Primary percutaneous coronary intervention; PI-PCI=Pharmaco-invasive percutaneous coronary intervention; STEMI=ST-segment elevation myocardial infarction; NSTEACS=non-ST-segment elevation acute coronary syndrome; CABG=coronary artery bypass graft; FMC=first medical contact

### 2.2 Definition of in-hospital outcomes

The primary efficacy outcomes were major adverse cardiac cerebrovascular events (MACCEs) that occurred during hospitalization. MACCEs were defined as cardiac death, non-fatal myocardial infarction, and acute stent thrombosis or ischemic stroke. The primary safety outcomes were defined as Thrombolysis in Myocardial Infarction (TIMI) major or minor bleeding during hospitalization.^9^ The net clinical outcomes (NET) were defined as a composite of the primary efficacy outcomes and the primary safety outcomes. The cardiac net clinical outcomes (cNET) were defined as MACCEs, new onset heart failure or new onset heart shock during hospitalization.

### 2.3 Statistical analysis

Continuous variables were shown as mean ± SD or median (interquartile range) according to different distributions and were compared using Student’s t-tests or Mann-Whitney U test. Categorical variables were presented as the number (percentage) and compared using chi-square test or Fisher exact tests. A linear-by-linear association trend test (Mantel-Haenszel test for trend) was used to assess trends of the ratios of different reperfusion therapy strategies. In-hospital clinical events were evaluated using multivariable logistic regression and adjusted for confounding factors that have been reported more than once as having an effect on outcomes. Candidate adjustment variables included age, female, previous myocardial infarction (MI), hypertension history, diabetes mellitus (DM) history, renal failure history, heart failure (HF) history, stroke history, atrial fibrillation (AF) history, dyslipidemia history, peripheral artery disease (PVD) history, previous PCI, previous coronary artery bypass grafting (CABG), anemia at admission, heart rate, systolic blood pressure (SBP), diastolic blood pressure (DBP), Killip class, hospital grade, length of hospital stay, low-density lipoprotein cholesterol (LDL-C), estimated glomerular filtration rate (eGFR). To consolidate the findings, we also carried out propensity score (PS) method in the study A logistic regression was performed to estimate PS, adjusting for the variables consistent with the multivariate analysis. We established PS-matched and inverse probability of treatment weighting (IPTW) cohort based on the PS score and evaluated the impact of different reperfusion strategies in STEMI. By this means we obtained a stabilized weight for each case of the study cohort, avoiding any extreme values that may result in unreliable outcomes. All tests were 2-sided, and a value of P < 0.05 was considered statistically significant. All statistical analyses were performed using Statistical Package for the Social Science (SPSS) 26.0 and R version 4.2.2.

## 3. Results

### 3.1 Situation of treatment strategies and clinical characteristics in STEMI

From November 2014 to December 2019, a total of 37733 patients were enrolled with a diagnosis of STEMI admitted within 48 hours after symptom onset and treated by thrombolysis or PCI.

Among them, 35019 patients received PPCI, 999 patients received thrombolysis and 1715 patients received PI-PCI. The ratios of different treatment strategies over the 5-year study period were shown in Figure 2. Between 2014 and 2019, The ratio of patients with STEMI received thrombolysis and PI-PCI increased over time while PPCI decreased.

**Figure 2.**
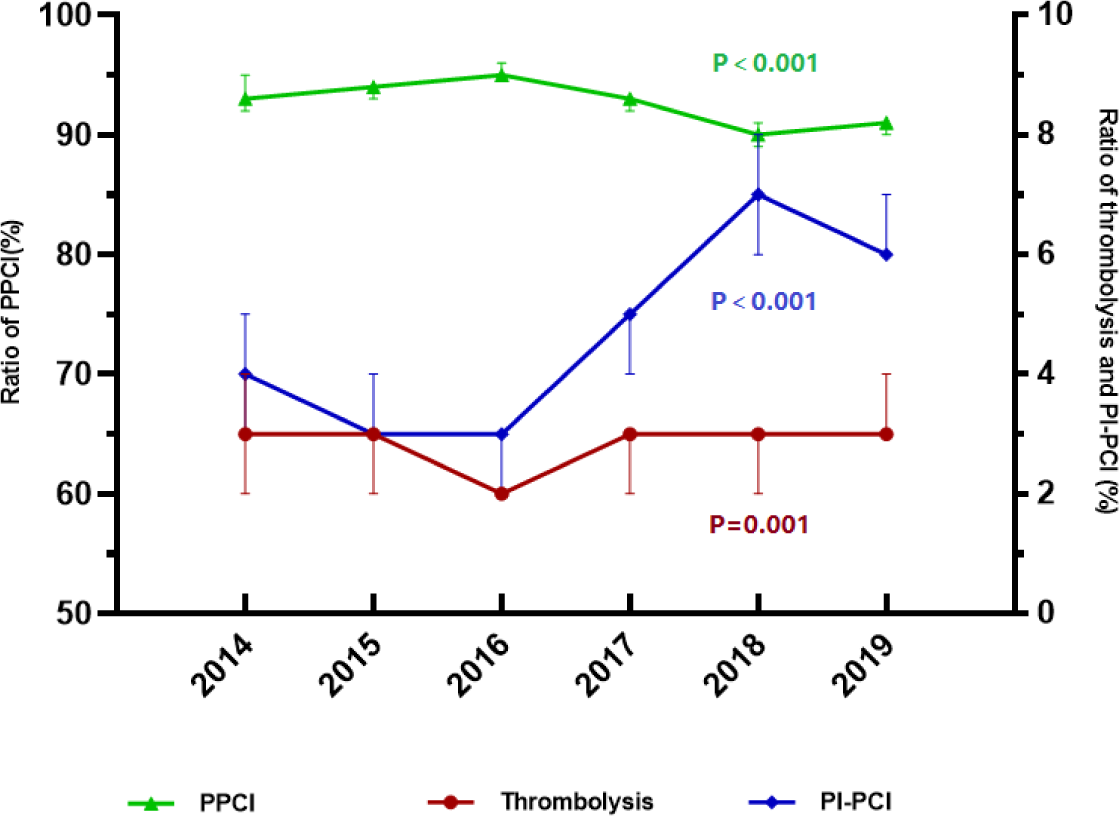
The ratios of PPCI, thrombolysis and PI-PCI from 2014 to 201

Compared with the PPCI group, the thrombolysis group had a higher proportion of previous MI, previous CABG, previous ischemic stroke history, Killip class I, statins at admission, anti-coagulation therapy, higher levels of blood pressure, LVEF and hemoglobin, and longer length of hospital stay. In contrast, the thrombolysis group had a lower proportion of previous PCI, DM, dyslipidemia, family history of coronary heart disease, tertiary hospital, β-blockers, warfarin, heart rate, the implantation of mechanical assist devices and temporary cardiac pacemaker during hospitalization, and lower levels of CK-MB, troponin I, troponin T, BNP and glucose (Table 1). Compared with the PPCI group, the PI-PCI group was younger, had a higher proportion of statins, higher levels of CK-MB, Troponin T and BNP, and longer length of hospital stay, lower levels of SBP and glucose, a lower proportion of female, tertiary hospital, previous PCI, hypertension, DM, stroke history, β-blockers, and the implantation of temporary cardiac pacemaker (Table 1).

**Table 1.** Baseline characteristics.

| Characteristics | PPCI (n=35019) | Thrombolysis (n=999) | PI-PCI (n=1715) | P value |
| --- | --- | --- | --- | --- |
| Age, y | 61.1±12.5 | 60.5±12.1 | 58.6±11.2<sup>b</sup> | <0.001 |
| Female, n (%) | 7047(20.1) | 196(19.6) | 299(17.4)<sup>b</sup> | 0.024 |
| Body mass index, kg/m<sup>2</sup> | 24.5±3.2 | 24.1±3.4 | 24.3±3.0<sup>b</sup> | 0.008 |
| Hospital grade, n (%) |  |  |  | <0.001 |
| Secondary hospital | 2471(7.1) | 313(31.3)<sup>a</sup> | 188(11.0)<sup>b</sup> |  |
| Tertiary hospital | 32548(92.9) | 686(68.7)<sup>a</sup> | 1527(89.0)<sup>b</sup> |  |
| Previous history, n (%) |  |  |  |  |
| Previous myocardial infarction | 1623(4.6) | 72(7.2)<sup>a</sup> | 75(4.4) | 0.001 |
| Previous PCI | 1715(4.9) | 30(3.0)<sup>a</sup> | 63(3.7)<sup>b</sup> | 0.002 |
| Previous CABG | 53(0.2) | 6(0.6)<sup>a</sup> | 3(0.2) | 0.003 |
| Hypertension | 16914(48.3) | 454(45.4) | 722(42.1)<sup>b</sup> | <0.001 |
| Dyslipidemia | 2062(5.9) | 37(3.7)<sup>a</sup> | 95(5.5) | 0.013 |
| Diabetes mellitus | 6677(19.1) | 163(16.3)<sup>a</sup> | 275(16.0)<sup>b</sup> | 0.001 |
| Chronic kidney disease | 271(0.8) | 13(1.3) | 8(0.5) | 0.057 |
| Atrial fibrillation | 440(1.3) | 15(1.5) | 13(0.8) | 0.143 |
| Heart failure | 153(0.4) | 9(0.9) | 10(0.6) | 0.072 |
| Cerebrovascular disease |  |  |  |  |
| Hemorrhagic stroke history | 241(0.7) | 3(0.3) | 2(0.1)<sup>b</sup> | 0.006 |
| Ischemic stroke history | 2163(6.2) | 85(8.5)<sup>a</sup> | 80(4.7)<sup>b</sup> | <0.001 |
| Peripheral arterial disease | 196(0.6) | 1(0.1) | 8(0.5) | 0.136 |
| COPD | 320(0.9) | 11(1.1) | 19(1.1) | 0.605 |
| Coronary heart disease family history | 1025(2.9) | 15(1.5)<sup>a</sup> | 42(2.4) | 0.016 |
| LVEF, % | 54.5±9.4 | 55.3±10.1<sup>a</sup> | 54.4±9.5 | 0.006 |
| Heart rate, beats/min | 78.1±15.1 | 75.6±17.4<sup>a</sup> | 76.3±15.6 | 0.008 |
| Systolic blood pressure, mmHg | 127.3±23.7 | 129.4±25.6<sup>a</sup> | 126.1±22.6<sup>b</sup> | <0.001 |
| Diastolic blood pressure, mmHg | 78.1±15.1 | 80.6±16.3<sup>a</sup> | 78.5±15.1 | 0.004 |
| Killip class, n (%) |  |  |  | 0.067 |
| I | 26456(75.5) | 780(78.1)<sup>a</sup> | 1280(74.6) |  |
| II or III | 7074(20.2) | 169(16.9) | 365(21.3) |  |
| IV | 1489(4.3) | 50(5.0) | 70(4.1) |  |
| Location of STEMI, n (%) |  |  |  |  |
| Anterior | 11998(34.3) | 321(32.1) | 603(35.2) | 0.269 |
| Inferior | 12387(35.4) | 367(36.7) | 547(31.9)<sup>b</sup> | 0.008 |
| Posterolateral | 987(2.8) | 26(2.6) | 35(2.0) | 0.151 |
| Others | 10726(30.6) | 325(28.4) | 580(33.8)<sup>b</sup> | 0.011 |
| Length of hospital stay, d | 9(7, 12) | 11(8, 14)<sup>a</sup> | 10(8, 14)<sup>b</sup> | <0.001 |
| Medications within 24 h, n (%) |  |  |  |  |
| Aspirin | 34214(97.7) | 980(98.1) | 1683(98.1) | 0.706 |
| P2Y12 inhibitors | 34284(97.9) | 975(97.6) | 1676(97.7) | 0.726 |
| ACEI/ARB | 15854(45.3) | 429(42.9) | 800(46.6) | 0.174 |
| β-blockers | 18512(52.9) | 489(48.9)<sup>a</sup> | 950(55.4)<sup>b</sup> | 0.005 |
| Statins | 33185(94.8) | 965(96.6)<sup>a</sup> | 1655(96.5)<sup>b</sup> | <0.001 |
| Warfarin | 186(0.5) | 1(0.1) | 6(0.3) | 0.107 |
| Clinical Laboratory |  |  |  |  |
| CK-MB, U/L | 49.3(15.8, 148.0) | 21.9(12.0, 62.1)<sup>a</sup> | 52.7(16.0, 187.7)<sup>b</sup> | <0.001 |
| Troponin T, ng/mL | 1.24(0.12, 6.36) | 0.37(0.05, 4.49)<sup>a</sup> | 2.20(0.35, 9.67)<sup>b</sup> | <0.001 |
| Troponin I, ng/mL | 2.60(0.17, 23.07) | 0.30(0.05, 2.70)<sup>a</sup> | 1.92(0.16, 26.30) | <0.001 |
| eGFR, ml/(min*1.73m<sup>2</sup>) | 91.2(74.0, 102.4) | 92.2(76.5, 102.8) | 93.6(77.6, 104.4)<sup>b</sup> | <0.001 |
| NT-proBNP, pg/ml | 402.0(110.0, 1313.0) | 419.7(100.0, 1239.3) | 473.4(128.9, 1266.0) | 0.339 |
| BNP, pg/ml | 105.1(33.0, 332.0) | 79.3(29.0, 236.0)<sup>a</sup> | 99.7(30.4, 309.1) | 0.025 |
| LDL-C, mmol/L | 2.82(2.23, 4.33) | 2.76(2.24, 3.39) | 2.71(2.14, 3.36) | <0.001 |
| Glucose, mmol/L | 6.27(5.23, 8.14) | 6.09(5.06, 7.90)<sup>a</sup> | 6.15(5.08, 7.83)<sup>b</sup> | <0.001 |
| Hemoglobin, g/L | 142.0(129.0, 153.2) | 143.0(132.0, 155.0)<sup>a</sup> | 142.0(130.0, 153.0) | 0.005 |
| HbA1C, % | 6.0(5.6, 7.1) | 5.9(5.5, 7.1) | 6.0(5.5, 7.0) | 0.078 |
| Anti-coagulation therapy, n (%) | 26591(75.9) | 920(92.1)<sup>a</sup> | 1304(76.0) | <0.001 |
| Mechanical circulatory assist device, n (%) | 397(1.1) | 0(0.0)<sup>a</sup> | 17(1.0) | <0.001 |
| Temporary cardiac pacemaker, n (%) | 369(1.1) | 2(0.2)<sup>a</sup> | 8(0.5)<sup>b</sup> | 0.002 |

### 3.2 In-hospital outcomes between PPCI group and thrombolysis group

After adjustment using the PS-matching method, baseline characteristics were well balanced (Supplemental Table 1). The incidence of all cause death (1.6% vs 2.8%, P=0.003), MACCEs (2.0% vs 3.6%, P<0.001), cardiac death, non-fatal MI, stroke, TIMI major bleedings (1.2% vs 2.2%, P=0.007), the net clinical outcomes, the cardiac net clinical outcomes, cardiac shock, cardiac arrest and new onset heart failure were significantly higher in the thrombolysis group than in the PPCI group, similar results were found after adjustment using the multivariate logistic analyses and PS-matching methods (Table 2). There was no significant difference in the incidence of TIMI minor bleedings between two groups (Table 2).

**Table 2.** In-hospital outcomes between PPCI group and thrombolysis group.

| Variables | PPCI | Thrombolysis | Unadjusted |  | Adjusted |  | PS-matched |  |
| --- | --- | --- | --- | --- | --- | --- | --- | --- |
|  |  |  | OR (95%CI) | P | OR (95% CI) | P | OR (95% CI) | P |
| All cause death, n (%) | 553(1.6) | 28(2.8) | 1.797(1.223-2.641) | 0.003 | 2.026(1.326-3.094) | 0.001 | 2.901(1.758-4.789) | < 0.001 |
| Cardiac death, n (%) | 511(1.5) | 26(2.6) | 1.805(1.211-2.690) | 0.004 | 2.111(1.361-3.276) | 0.001 | 2.919(1.734-4.917) | < 0.001 |
| MACCEs, n (%) | 694(2.0) | 36(3.6) | 1.849(1.315-2.601) | < 0.001 | 1.939(1.341-2.804) | 0.001 | 2.539(1.652-3.903) | < 0.001 |
| Non-fatal MI, n (%) | 108(0.3) | 11(1.1) | 3.599(1.929-6.714) | < 0.001 | 3.368(1.739-6.523) | 0.001 | 3.329(1.487-7.454) | 0.003 |
| Stroke, n (%) | 145(0.4) | 14(1.4) | 3.418(1.968-5.938) | < 0.001 | 3.810(2.112-6.874) | < 0.001 | 3.682(1.771-7.654) | < 0.001 |
| Cardiac shock, n (%) | 1090(3.1) | 70(7.0) | 2.345(1.826-3.013) | < 0.001 | 2.800(2.072-3.785) | < 0.001 | 2.473(1.814-3.371) | < 0.001 |
| Cardiac arrest, n (%) | 607(1.7) | 30(3.0) | 1.755(1.210-2.546) | 0.003 | 1.562(1.045-2.336) | 0.030 | 2.165(1.380-3.397) | 0.001 |
| Heart failure, n (%) | 4308(12.3) | 167(16.7) | 1.431(1.208-1.695) | < 0.001 | 1.786(1.437-2.219) | < 0.001 | 1.418(1.168-1.721) | < 0.001 |
| TIMI major and minor bleedings, n (%) | 2177(6.2) | 66(6.6) | 1.067(0.828-1.375) | 0.615 | 0.996(0.767-1.293) | 0.975 | 1.055(0.796-1.398) | 0.708 |
| TIMI major bleedings, n (%) | 431(1.2) | 22(2.2) | 1.807(1.172-2.787) | 0.007 | 1.993(1.275-3.116) | 0.002 | 1.807(1.086-3.009) | 0.023 |
| TIMI minor bleedings, n (%) | 1746(5.0) | 44(4.4) | 0.878(0.647-1.192) | 0.405 | 0.884(0.647-1.208) | 0.222 | 0.866(0.620-1.210) | 0.439 |
| NET, n (%) | 2766(7.9) | 99(9.9) | 1.283(1.039-1.584) | 0.021 | 1.325(1.064-1.651) | 0.012 | 1.330(1.046-1.691) | 0.020 |
| cNET, n (%) | 4958(14.2) | 216(21.6) | 1.673(1.434-1.950) | < 0.001 | 2.028(1.727-2.511) | < 0.001 | 1.660(1.390-1.982) | < 0.001 |

### 3.3 In-hospital outcomes between PPCI group and PI-PCI group

After adjustment using the PS-matched method, baseline characteristics were well balanced (Supplemental Table 2). The incidence of MACCEs (2.0% vs 0.8%, P=0.001), all cause death and cardiac death were significantly lower in the PI-PCI group and similar results were found after adjustment using the multivariate logistic analyses and the PS-matching methods. The risk of new onset heart failure and the cardiac net clinical outcomes were higher in PI-PCI group and the difference were statistically significant after adjustment using the multivariate logistic analyses. There was no significant difference in the incidence of outcomes including stroke, cardiac arrest, cardiac shock, TIMI major or TIMI major bleedings and the net clinical outcomes between two groups. (Table 3).

**Table 3.** In-hospital outcomes between PPCI group and PI-PCI group.

| Variables | PPCI | PI-PCI | Unadjusted |  | Adjusted |  | PS-matched |  |
| --- | --- | --- | --- | --- | --- | --- | --- | --- |
|  |  |  | OR 95% CI | P | OR 95% CI | P | OR 95% CI | P |
| All cause death, n (%) | 553(1.6) | 8(0.4) | 0.292(0.145-0.588) | 0.001 | 0.354(0.174-0.722) | 0.004 | 0.408(0.196-0.846) | 0.016 |
| Cardiac death, n (%) | 511(1.5) | 7(0.4) | 0.277(0.131-0.584) | 0.001 | 0.340(0.159-0.726) | 0.005 | 0.404(0.185-0.881) | 0.023 |
| MACCEs, n (%) | 694(2.0) | 14(0.8) | 0.407(0.239-0.693) | 0.001 | 0.468(0.273-0.802) | 0.006 | 0.481(0.275-0.840) | 0.010 |
| Stent thrombosis, n (%) | 79(0.2) | 3(0.2) | 0.775(0.244-2.457) | 0.665 | 0.749(0.235-2.383) | 0.624 | 0.514(0.154-1.1715) | 0.279 |
| Non-fatal MI, n (%) | 108(0.3) | 6(0.3) | 1.135(0.498-2.586) | 0.763 | 1.195(0.521-2.742) | 0.674 | 1.077(0.436-2.661) | 0.872 |
| Stroke, n (%) | 145(0.4) | 4(0.2) | 0.562(0.208-1.520) | 0.257 | 0.624(0.230-1.698) | 0.356 | 0.526(0.185-1.494) | 0.227 |
| Cardiac shock, n (%) | 1090(3.1) | 52(3.0) | 0.973(0.734-1.291) | 0.851 | 1.093(0.801-1.491) | 0.575 | 1.084(0.794-1.479) | 0.614 |
| Cardiac arrest, n (%) | 607(1.7) | 22(1.3) | 0.737(0.480-1.130) | 0.162 | 0.824(0.532-1.277) | 0.387 | 0.923(0.579-1.473) | 0.738 |
| Heart failure, n (%) | 4308(12.3) | 233(13.6) | 1.121(0.973-1.291) | 0.115 | 1.316(1.101-1.573) | 0.003 | 1.137(0.972-1.329) | 0.108 |
| TIMI major and minor bleedings, n (%) | 2177(6.2) | 126(7.4) | 1.196(0.993-1.442) | 0.060 | 1.119(0.925-1.355) | 0.247 | 1.106(0.901-1.358) | 0.333 |
| TIMI major bleedings, n (%) | 431(1.2) | 29(1.7) | 1.380(0.945-2.017) | 0.096 | 1.450(0.990-2.124) | 0.057 | 1.222(0.803-1.860) | 0.349 |
| TIMI minor bleedings, n (%) | 1746(5.0) | 97(5.7) | 1.142(0.926-1.410) | 0.215 | 1.199(0.970-1.483) | 0.093 | 1.071(0.850-1.349) | 0.559 |
| NET, n (%) | 2766(7.9) | 139(8.1) | 1.028(0.861-1.228) | 0.757 | 1.104(0.922-1.323) | 0.283 | 1.012(0.833-1.229) | 0.904 |
| cNET, n (%) | 4958(14.2) | 262(15.3) | 1.093(0.955-1.251) | 0.195 | 1.233(1.046-1.453) | 0.012 | 1.099(0.947-1.274) | 0.214 |

OR=odds ratio; CI=confidence interval; MACCEs=major adverse cardiac cerebrovascular events; TIMI=Thrombolysis in Myocardial Infarction; MI=myocardial infarction; PPCI=primary percutaneous coronary intervention; PI-PCI=pharmaco-invasive primary percutaneous coronary; PS=propensity score; NET=net clinical outcomes; cNET=cardiac net clinical outcomes

### 3.4 Subgroup analysis

#### 3.4.1 Comparison of outcomes between PPCI with PI-PCI groups in the time from onset to FMC

In the subgroup analyses, the time from onset to FMC ≥ 3h, 22413 patients received PPCI and 1211 patients received PI-PCI. After adjustment using the PS-matching method, baseline characteristics were well balanced (Supplemental Table 3).

The incidence of all cause death (0.2% vs 1.6%, P =0.001) and cardiac death (0.2% vs 1.5%, P =0.002) were lower in the PI-PCI group than in the PPCI group in all models. The incidence of MACCEs (0.7% vs 1.9%, P =0.003) was lower in the PI-PCI group than in the PPCI group in unadjusted and adjusted models. The risk of TIMI major and minor bleedings was significantly higher in the PI-PCI group in all cohorts. The ratio of new onset heart failure, the net clinical outcomes and the cardiac net clinical outcomes were significantly higher in PI-PCI group in the multivariate logistic analyses cohort. There was no significant difference in outcomes including non-fatal MI, stent thrombosis,cardiac shock,cardiac arrest and stroke (Table 4).

**Table 4.** In-hospital outcomes between PPCI group and PI-PCI group in the time from onset to FMC ≥ 3h.

| Variables | PPCI<br>(n=22413) | PI-PCI<br>(n=1211) | Unadjusted |  | Adjusted |  | PS-matched |  |
| --- | --- | --- | --- | --- | --- | --- | --- | --- |
|  |  |  | OR 95% CI | P | OR 95% CI | P | OR 95% CI | P |
| All cause death, n (%) | 360(1.6) | 3(0.2) | 0.152(0.049-0.475) | 0.001 | 0.177(0.056-0.558) | 0.003 | 0.259(0.080-0.836) | 0.024 |
| Cardiac death, n (%) | 342(1.5) | 3(0.2) | 0.180(0.051-0.500) | 0.002 | 0.189(0.060-0.597) | 0.005 | 0.278(0.086-0.899) | 0.032 |
| MACCE, n (%) | 429(1.9) | 8(0.7) | 0.341(0.169-0.687) | 0.003 | 0.388(0.190-0.793) | 0.009 | 0.511(0.244-1.070) | 0.075 |
| Stent thrombosis, n (%) | 41(0.2) | 1(0.1) | 0.451(0.062-3.281) | 0.432 | 0.436(0.059-3.206) | 0.415 | 0.559(0.069-4.550) | 0.587 |
| Non-fatal MI, n (%) | 55(0.2) | 4(0.3) | 1.347(0.487-3.723) | 0.566 | 1.453(0.520-4.063) | 0.476 | 1.962(0.590-6.527) | 0.272 |
| Stroke, n (%) | 71(0.3) | 3(0.2) | 0.781(0.246-2.485) | 0.676 | 0.836(0.260-2.694) | 0.765 | 1.056(0.737-1.512) | 0.555 |
| Cardiac shock, n (%) | 707(3.2) | 40(3.3) | 1.049(0.759-1.450) | 0.773 | 1.166(0.814-1.670) | 0.401 | 1.056(0.737-1.512) | 0.768 |
| Cardiac arrest, n (%) | 384(1.7) | 16(1.3) | 0.768(0.464-1.271) | 0.304 | 0.862(0.515-1.445) | 0.574 | 1.208(0.687-2.124) | 0.511 |
| Heart failure, n (%) | 2760(12.3) | 166(13.7) | 1.131(0.956-1.339) | 0.152 | 1.343(1.086-1.660) | 0.006 | 1.097(0.911-1.321) | 0.331 |
| TIMI major and minor bleedings, n (%) | 1412(6.3) | 103(8.5) | 1.383(1.122-1.704) | 0.002 | 1.273(1.028-1.578) | 0.027 | 1.277(1.013-1.610) | 0.039 |
| TIMI major bleedings, n (%) | 268(1.2) | 20(1.7) | 1.388(0.878-2.194) | 0.161 | 1.467(0.925-2.328) | 0.104 | 1.248(0.752-2.072) | 0.392 |
| TIMI minor bleedings, n (%) | 1144(5.1) | 83(6.7) | 1.368(1.086-1.723) | 0.008 | 1.431(1.133-1.807) | 0.003 | 1.274(0.987-1.646) | 0.063 |
| NET, n (%) | 1782(8.0) | 110(9.1) | 1.157(0.945-1.415) | 0.158 | 1.247(1.015-1.532) | 0.036 | 1.171(0.937-1.463) | 0.165 |
| cNET, n (%) | 3161(14.1) | 190(15.7) | 1.133(0.966-1.329) | 0.124 | 1.319(1.086-1.601) | 0.005 | 1.110(0.931-1.323) | 0.245 |

OR= odds ratio; CI= confidence interval; MACCEs=major adverse cardiac cerebrovascular events; TIMI=Thrombolysis in Myocardial Infarction; MI=myocardial infarction; PS=propensity score; PPCI=primary percutaneous coronary intervention; PI-PCI=Pharmaco-invasive percutaneous coronary intervention; FMC=first medical contact; NET=net clinical outcomes;cNET=cardiac net clinical outcomes

#### 3.4.2 Comparison of outcomes between PPCI with PI-PCI groups according to the reperfusion timing

Baseline characteristics are summarized in Supplemental Table 4. Based on the reperfusion timing, we divided the PPCI group into the Timely PPCI (time from FMC to wire crossing of the infarction related artery ≤ 120 mins) and Late PPCI (time from FMC to wire crossing of the infarction related artery > 120 mins) groups; The PI-PCI is divided into scheduled PCI (PCI was scheduled within 24 hours after successful thrombolysis) and rescue PCI (PCI was performed immediately after thrombolysis failure) (Supplemental Table 5).

Compared with timely PPCI group, the incidence of all cause death was significantly lower in scheduled PCI group in all model. The incidence of MACCEs was statistically significant lower in the scheduled PCI group in the adjusted model. However, the incidence of heart failure was significantly higher in the scheduled PCI group in all models. The incidence of TIMI major or minor bleedings and the cardiac net clinical outcomes was statistically significant lower in scheduled PCI group in the unadjusted and adjusted models (Figure 3 A). Compared with late PPCI group, the incidence of all cause death, MACCEs were significantly lower in scheduled PCI group in all models and the incidence of heart failure was significantly higher in the scheduled PCI group in the unadjusted and adjusted models (Figure 3 B). Compared with timely PPCI, the ratio of heart failure was statistically significant higher in the rescue PCI group in the adjusted and PS-matched models and the ratio of the cardiac net clinical outcomes was higher in the rescue PCI group in the PS-matched model.(Figure 3 C). There was no significant difference in all outcomes in all models between rescue PCI group and late PPCI group (Figure 3 D).

For further analysis, according to the time to PCI after thrombolysis in scheduled PCI group, we divided the patients into 3 groups: scheduled PCI ≤ 24h group, scheduled PCI during 24h to 7d group, and scheduled PCI>7d group. Compared with scheduled PCI ≤ 24h group, the rate of TIMI major or minor bleedings and the net clinical outcomes were lower in scheduled PCI during 24h to 7d group in all models, and the rate of heart failure was also lower in scheduled PCI during 24h to 7d group in unadjusted model (Figure 3 E). In addition, compared with scheduled PCI ≤ 24h group, the scheduled PCI >7d group had lower risk of heart failure in all models, and lower cardiac net clinical outcomes in IPTW model (Figure 3 F).

**Figure 3.**
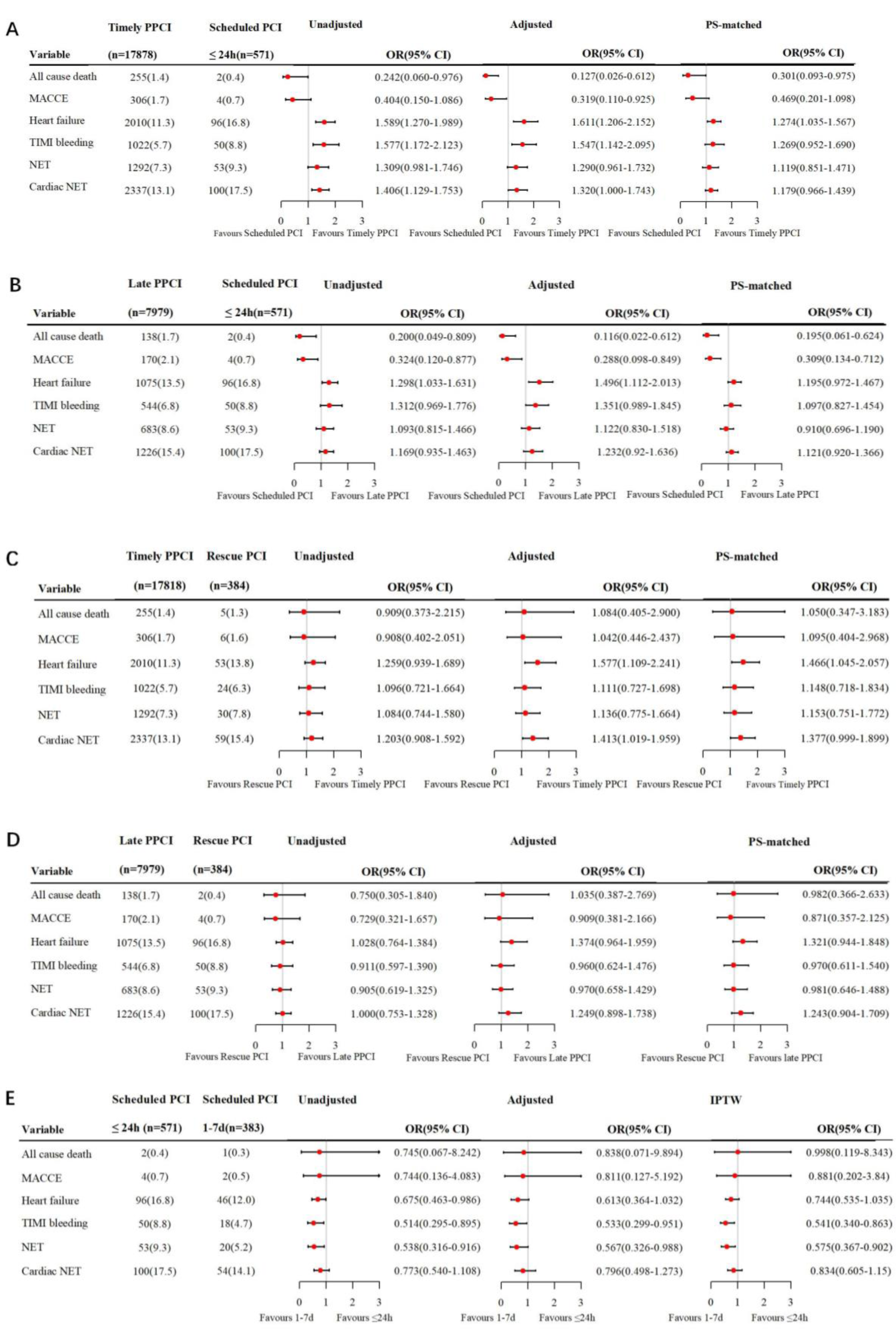
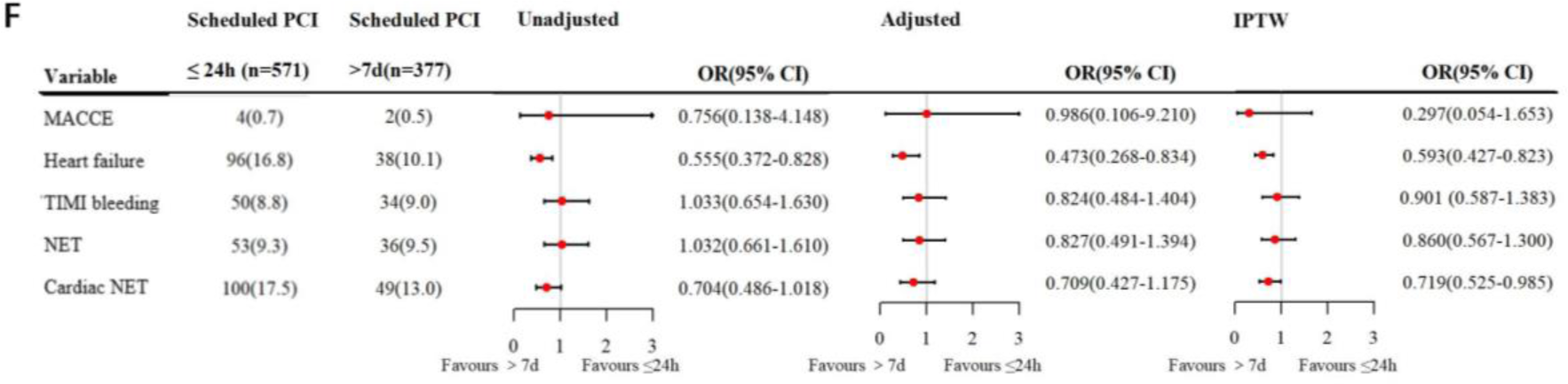
Main outcomes in subgroup analysis between PPCI with PI-PCI groups according to the reperfusion timing

Based on the reperfusion timing, we divided the PPCI group into the Timely PPCI (time from FMC to the wire crossing of the infarction related artery ≤ 120min) and Late PPCI (time from FMC to the wire crossing of the infarction related artery > 120min) groups; The PI-PCI is divided into scheduled PCI (PCI was scheduled within 24 hours after successful thrombolysis) and rescue PCI (PCI was performed immediately after thrombolysis failure); According to the time to PCI after thrombolysis in scheduled PCI group, we divided the patients into 3 groups: scheduled PCI ≤ 24h group, scheduled PCI during 1d to 7d group and scheduled PCI > 7d group.

A. Comparing scheduled PCI with timely PPCI in main outcomes by unadjusted model, adjusted model and PS-matched model;
B. Comparing scheduled PCI with late PPCI in main outcomes by unadjusted model, adjusted model and PS-matched model;
C. Comparing rescue PCI with timely PPCI in main outcomes by unadjusted model, adjusted model and PS-matched model;
D. Comparing rescue PCI with late PPCI in main outcomes by unadjusted model, adjusted model and PS-matched model;
E. Compared scheduled PCI ≤24h with scheduled PCI during 1∼7d in main outcomes used by unadjusted model, adjusted model and IPTW model;
F. Comparing scheduled PCI ≤ 24h with scheduled PCI > 7d in main outcomes by unadjusted model, adjusted model and IPTW model.

OR=odds ratio; CI=confidence interval; MACCEs=major adverse cardiac cerebrovascular events; TIMI=Thrombolysis in Myocardial Infarction; PCI=primary percutaneous coronary intervention; PS=propensity score; IPTW=inverse probability of treatment weighting; PPCI=primary percutaneous coronary; FMC=first medical contact; PI-PCI=Pharmaco-invasive percutaneous coronary intervention

## 4. Discussion

In our study, nearly 10% patients with STEMI received thrombolysis, in which 2.6% of the patients received thrombolysis alone. Consistent with the previous reports, our study showed that patients with STEMI were more male and had comorbidities, such as hypertension, DM and ischemic diseases (including previous coronary heart diseases, ischemic stroke, and PAD).^10, 11^ Although PPCI is the reperfusion strategy of choice in STEMI patients when performed by an experienced team in a timely manner, thrombolysis therapy is still more feasible and often the only available, even the mainstay reperfusion strategy for STEMI in resource-poor settings without advanced technology or access to specialized care, especially in county-level hospitals.10,12

Thrombolytic therapy can improve the perfusion of the coronary artery and the microcirculation system. The following points should be noted in the thrombolysis. Firstly, the effect of thrombolysis has a clear time-dependence, both clinical guidelines and previous studies indicated that onset time ≤3h was independent predictors of successful thrombolysis.^7,11,13–17^ However, the proportion of patients from the time of onset to thrombolysis ≤ 3h in China was only 27.9%, while the proportion in the same period (2014) was as high as 54% in the United States of America, indicating that the pre-hospital delay of thrombolysis in STEMI patients in China is still serious problem.^18^ Secondly, the use of fibrin-specific thrombolytic agents was also a key factor in the success of thrombolysis therapy. In our study, the proportion of patients used non-fibrin-specific agents in thrombolysis group was 32.1%, which may lead to the poor efficacy of thrombolytic therapy. At last, hemorrhage is the most common complication of thrombolysis therapy, especially intracranial hemorrhage (0.9-1%). In our study, the rate of TIMI major bleedings was also significantly higher in thrombolysis group. The CAMI study showed that compared with urokinase, alteplase and reteplase had a higher rate of thrombolytic success, but more in-hospital bleeding events.^15^ Previous research found that prourokinase, as a novel and fibrin-specific anticoagulant, not only effectively recanalized blood vessels and improve the cardiac function of patients with STEMI, but also had very low rate of bleeding complications, which may provide an alternative and appropriate thrombolytic drug option for STEMI patients.^19, 20^

Besides, this study showed that patients receiving thrombolysis alone had worse in-hospital ischemic outcomes. Previous studies have shown that plaque rupture and plaque erosion are the 2 most common underlying mechanisms for sudden cardiac death and acute coronary syndrome. It has been well known that the necrotic core is 6 times more thrombogenic than all other plaque components.^21,22,23^ Confirmed by optical coherence tomography(OCT), residual thrombus burden 1 day after fibrinolysis was greater in a rupture compared with erosion in patients with successful fibrinolysis for STEMI.^24^ The advantage of the thrombolytic therapy is to dissolve the large, medium, and microthrombus in coronary arteries, which can improve the perfusion of the coronary artery and the microcirculation system. However, the reoccurrence of myocardial ischemia after thrombolytic therapy is common. Therefore, thrombolysis therapy alone is not sufficient for patients with STEMI.

Previous studies showed that PI-PCI was not inferior to PPCI, and even better in follow-up in ischemic or mortality events. In the STREAM study, the composite end point of death, cardiac shock, congestive HF, and reinfarction was numerically but non-statistically significant lower in the pharmaco-invasive (PI) arm at 30 days.^25^ The EARLY-MYO Trial indicated that a PI strategy with half-dose alteplase and timely PPCI offered more complete epicardial and myocardial reperfusion compared with PPCI in patients with STEMI presenting ≤ 6 hours after symptom onset but with an expected PCI-related delay in China.^26^ The STREAM-2 study, enrolling in patients aged ≥ 60 years with STEMI, showed that there were no differences in lytic versus PPCI patients, either in symptom onset to start of reperfusion time, or in final TIMI 3 flow after PCI or last angiography, despite a higher baseline TIMI 0/1 flow prior to PCI in the intervention group.^27^ The results of this study indicated that PI-PCI was superior to PPCI in mortality and MACCEs during hospitalization,also in the subgroup of the time from onset to FMC ≥ 3h. However, PI-PCI had higher risk of TIMI major or minor bleedings than PPCI in the subgroup of the time from onset to FMC ≥ 3h. A Norway Study showed similar results in patients who did not have PPCI performed within 120 mins.^28^An increased risk of bleeding may be caused by simultaneous application of thrombolytic and anticoagulant drugs in PI-PCI strategy. In our study, the incidence of heart failure was higher in the PI-PCI group. Movement of patients during transport increase the cardiac burden and more early reperfusion of PI-PCI increase the occurrence of malignant arrhythmia, which may increase the occurrence of in-hospital new onset HF in PI-PCI.

A meta-analysis compared pharmacoinvasive therapy (PIT, defined as administration of thrombolytic drugs followed by immediate PCI only in case of failed thrombolysis) with PPCI, showing that PIT significantly decreased short-term mortality [OR=1.46(1.08 to 1.96), I2=0%, P=0.01] in those studies with a 200 mins of symptom-onset-to-device time.^29^ Jamal *et al.* found that patients who underwent late PPCI had higher mortality rates than those underwent a PI strategy.^30^ In our study, compared with timely PPCI and late PPCI, scheduled PCI group was better in in-hospital mortality and MACCEs, which indicated early thrombolytic therapy may further shorten the myocardial ischemia time and reduce the ischemic events. The rate of heart failure was higher in the scheduled PCI, which may be caused by the worse condition of the scheduled PCI group, especially in the ratio of Killip II-IV class.

European Society of Cardiology (ESC) and Chinese guidelines recommend a door-to-balloon time of ≤120 minutes.^7,11^ American College of Cardiology and American Heart Association (ACC/AHA) guidelines recommend a door-to-balloon time of <90 minutes.^14^ There are various conditions, including occupancy of cardiac catheterization laboratory, will of patients and their families, and unsuitable clinical conditions of patient, inevitably increase the time to revascularization in the clinic and leading to increased risk of in-hospital mortality. At present, the total ischemic time rather than door-to-balloon time is the principal determinant of outcomes in STEMI. PI-PCI is a possible strategy better than PPCI in patients requiring transfer. PI-PCI can shorten the time of myocardial ischemia, significantly reduce the occurrence of no-reflow, markedly increase the proportion of TIMI 3 flow, expand the subsequent PCI time by early thrombolysis, and reduce the mortality rate.^31^ Therefore, for patients who cannot perform timely PCI, the reperfusion strategy of PI-PCI is relatively effective and safe. The factors needs to be considered including symptom-onset to first medical contact time, expected time of transfer to a PCI-capable hospital, and patients risk factors. However, it is notable that PI-PCI may increase the risk of bleeding during hospitalization. In suitable patients, PI-PCI should be evaluated in large clinical trials to assess its clinical and safety outcomes.

Our study also compared the in-hospital efficacy of performing PCI at different times after successful thrombolysis. Compared with scheduled PCI ≤ 24h group, the scheduled PCI during 24h to 7d group had lower risk of TIMI major or minor bleedings,which may be caused by simultaneous application of thrombolytic drugs and anticoagulant drugs within a short time.However, the scheduled PCI >7d group had the similar risk of bleedings with scheduled PCI ≤ 24h group, which may show that the prolonged using of anticoagulants achieved a high bleeding risk similar to the simultaneous application of anticoagulants and thrombolytic drugs.In addition, the proportion of receiving anti-coagulation therapy in the scheduled PCI during 24h to 7d group was lower than both the scheduled PCI ≤ 24h group and the scheduled PCI >7d group, which may another reason for the low incidence of bleeding.Besides, compared with scheduled PCI ≤ 24h group, the scheduled PCI >7d group has lower risk of heart failure. This study is a retrospective study, the treatment strategy of patients depends on the judgment of the doctor, and patients with serious clinical conditions are more inclined toward early revascularization. In the baseline data, the ratio of Killip II-IV class in scheduled PCI >7d group was the lowest (20.4%) among all the subgroups, which supported the above speculation. Therefore, it is feasible to appropriately prolong the scheduled PCI time in STEMI patients, accompanied by more stable clinical conditions, such as hemodynamic stability, stable symptoms, etc.

## 5. Limitation

First, this was a retrospective observational study, so we could not rule out unmeasured potential confounders. Second, because the participating hospitals were not randomly selected, which may not reflect the clinical situation of all hospitals in China. Finally, this study only analyzed in-hospital outcomes and did not include long-term follow-up.

## 6. Conclusions

In STEMI patients who could not perform timely PPCI, PI-PCI is feasible, including rescue PCI,which can reduce the rate of MACCEs and mortality during hospitalization.But the increased risk of bleedings also should be concerned.In scheduled PCI after successful thrombolysis, appropriate extension the time window of scheduled PCI can be considered under stable clinical conditions.

## Data Availability

Due to the nature of this research, participants of this study did not agree for their data to be shared publicly, so supporting data is not available.

## Acknowledgement

The authors thank Dr Jun Liu (Department of Epidemiology, Beijing Anzhen Hospital, Capital Medical University, Beijing Institute of Heart, Lung and Blood Vessel Diseases, Beijing, China) for her technical assistance. We acknowledge all participating hospitals for their contributions to CCC-ACS Project (Supplemental Table 6).

---

**Supplemental Table 1.**
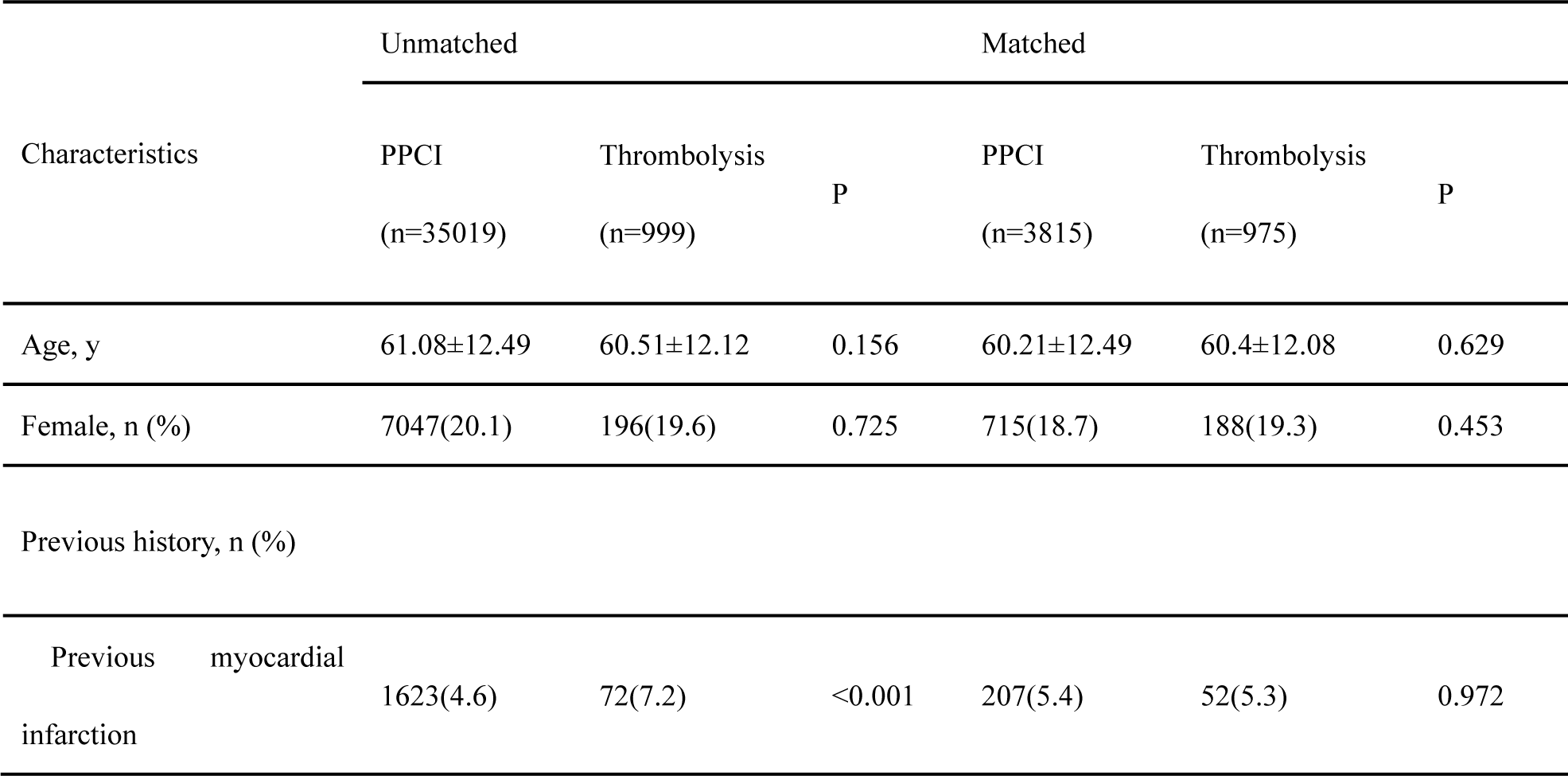
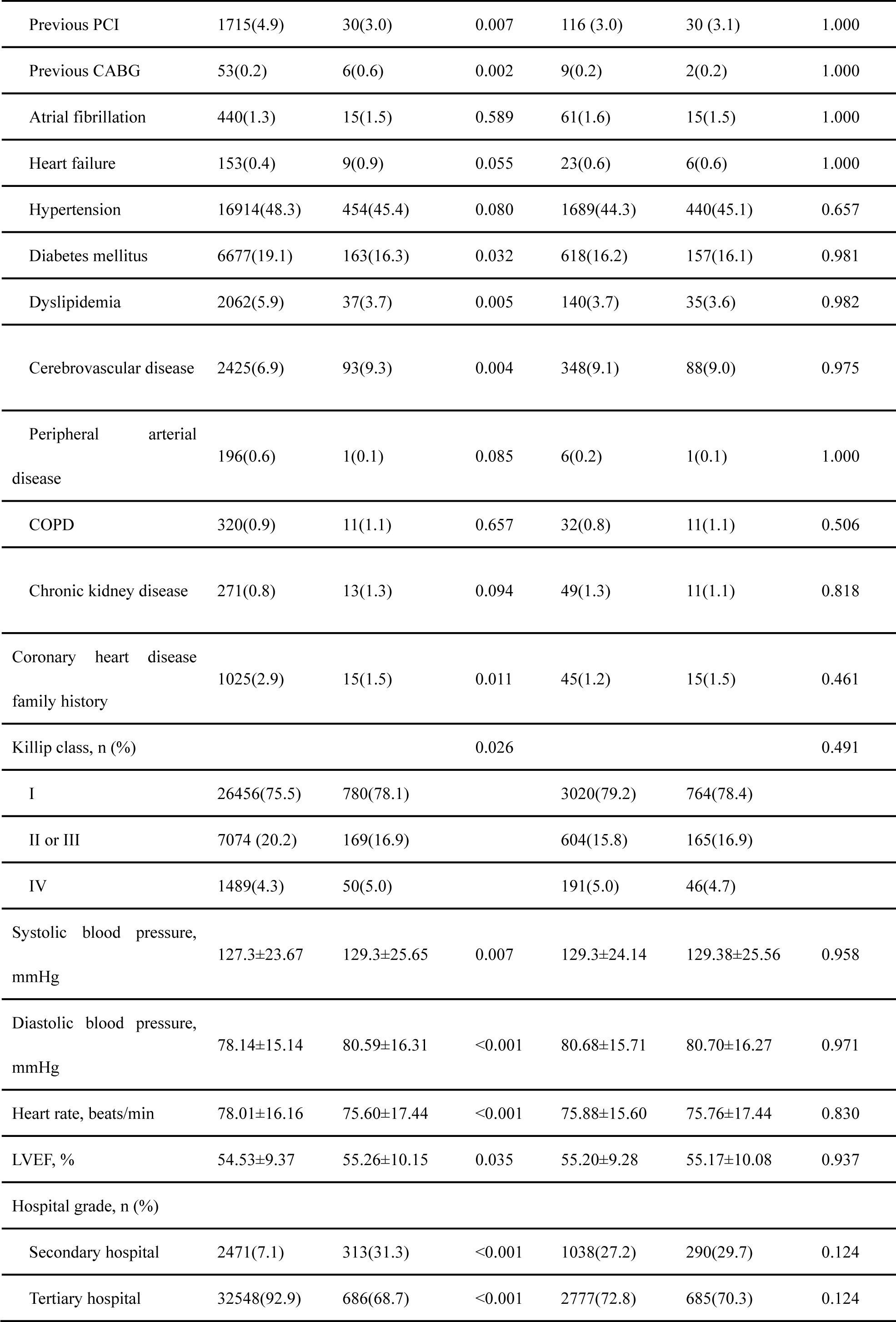
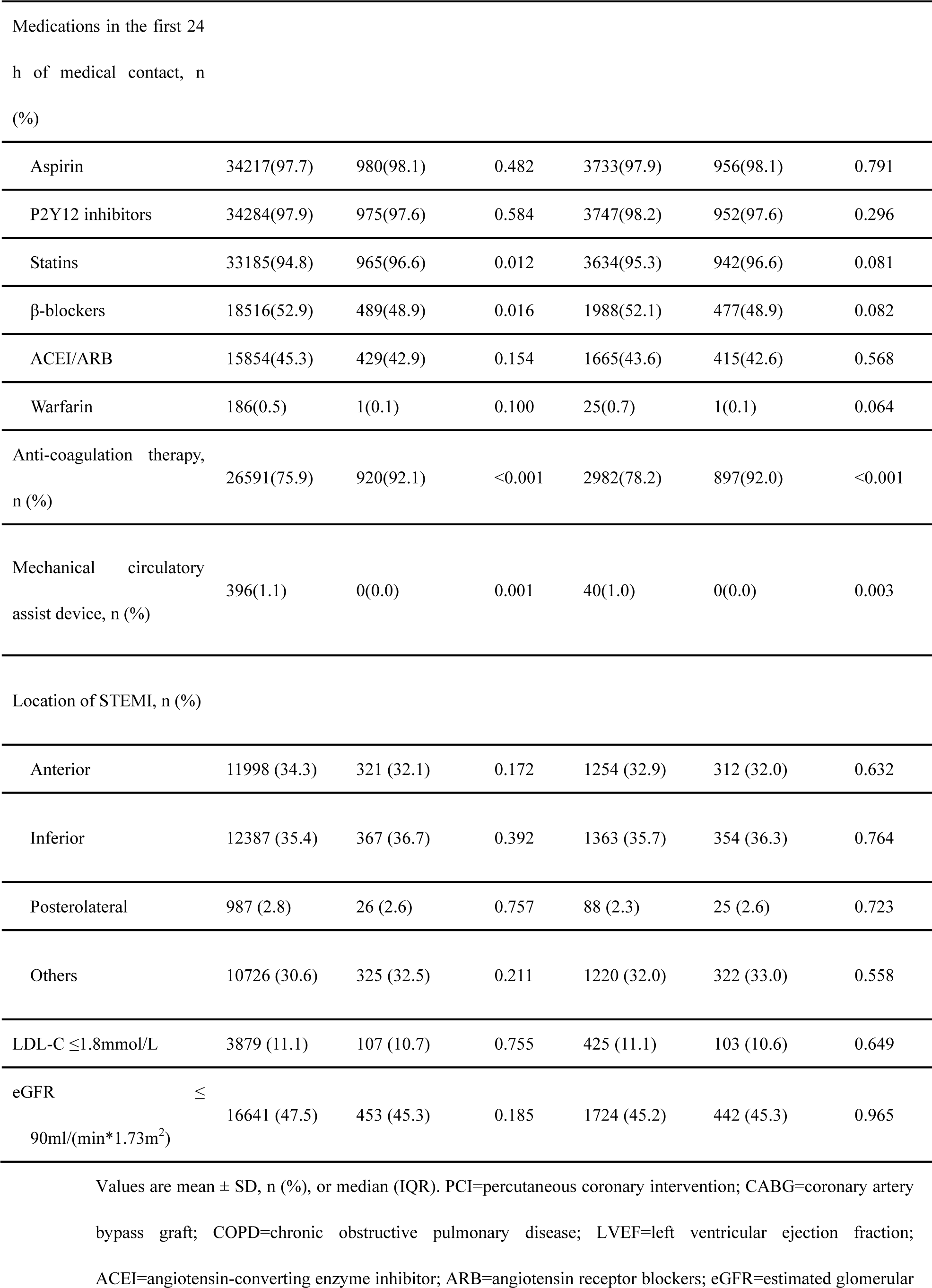
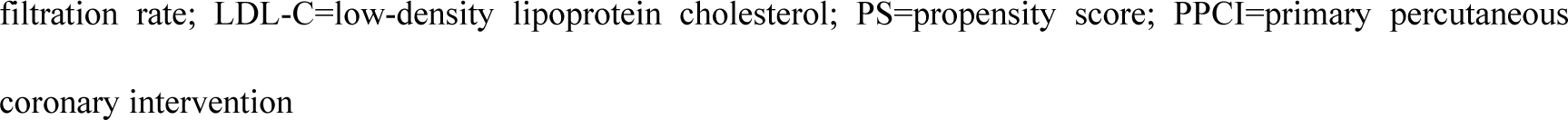
Comparison of baseline characteristics between PPCI and Thrombolysis groups in the whole, PS-matched and IPTW cohorts.

**Supplemental Table 2.**
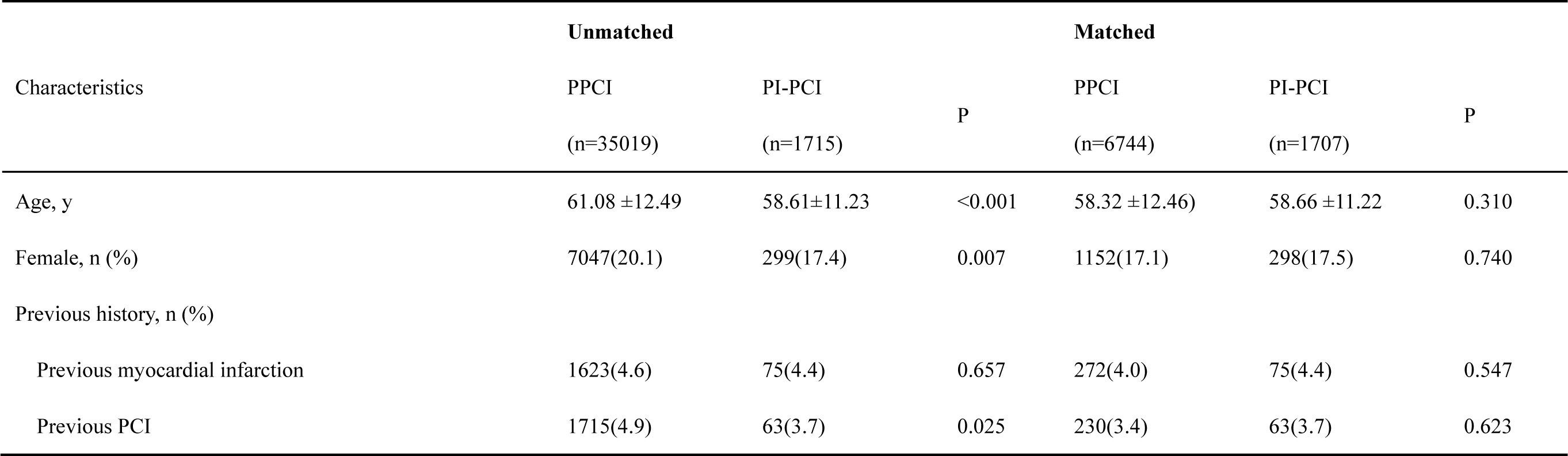
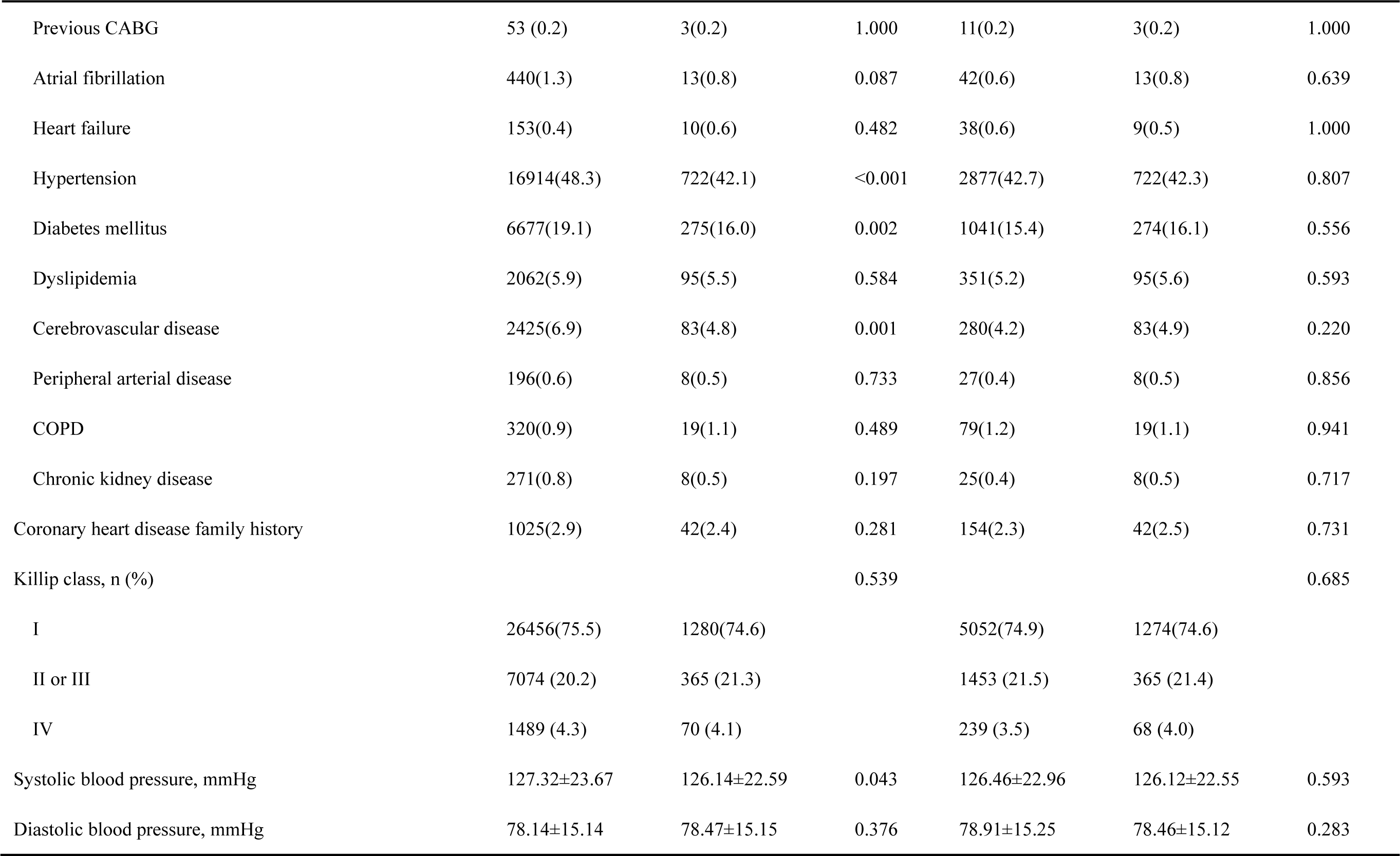
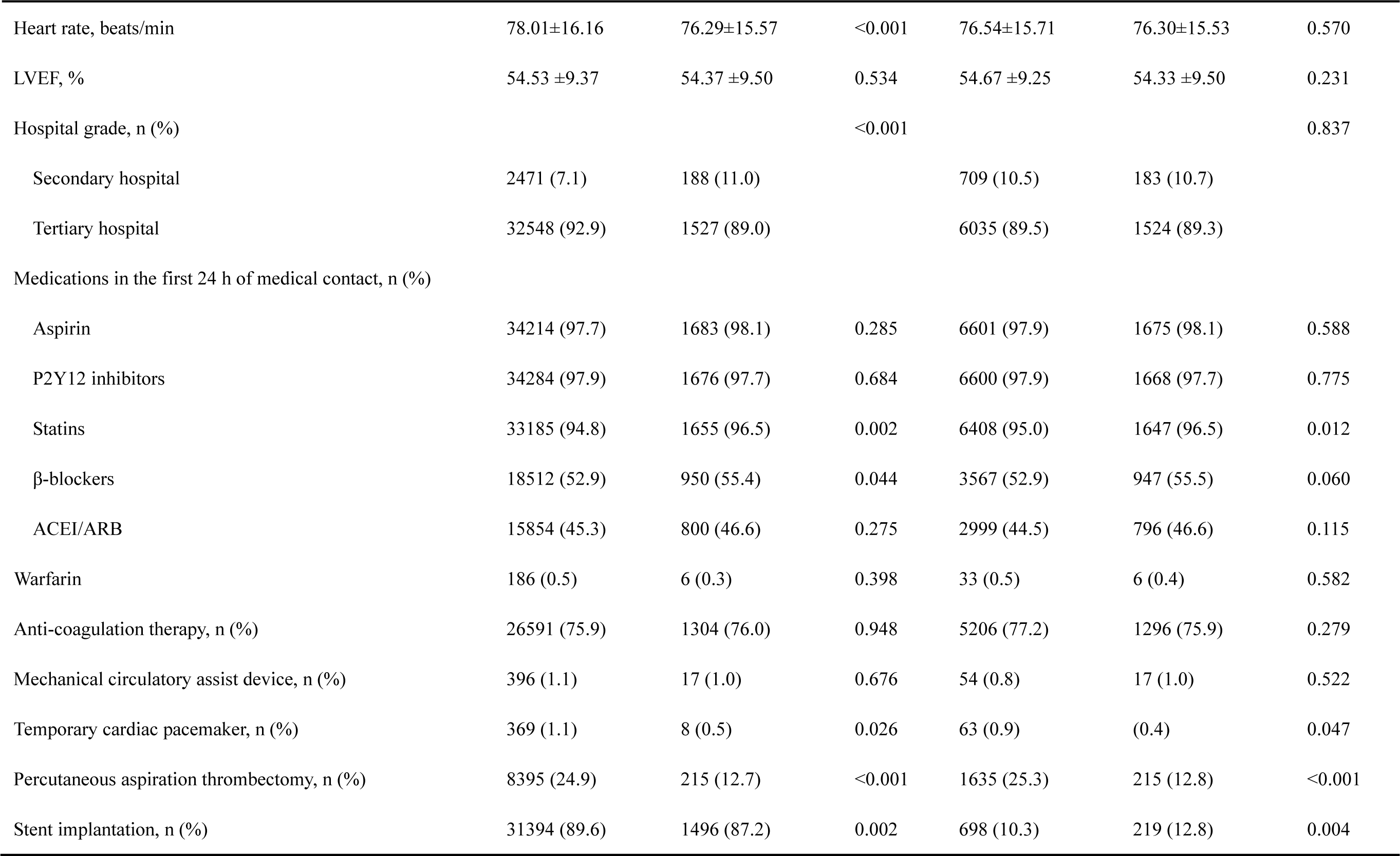
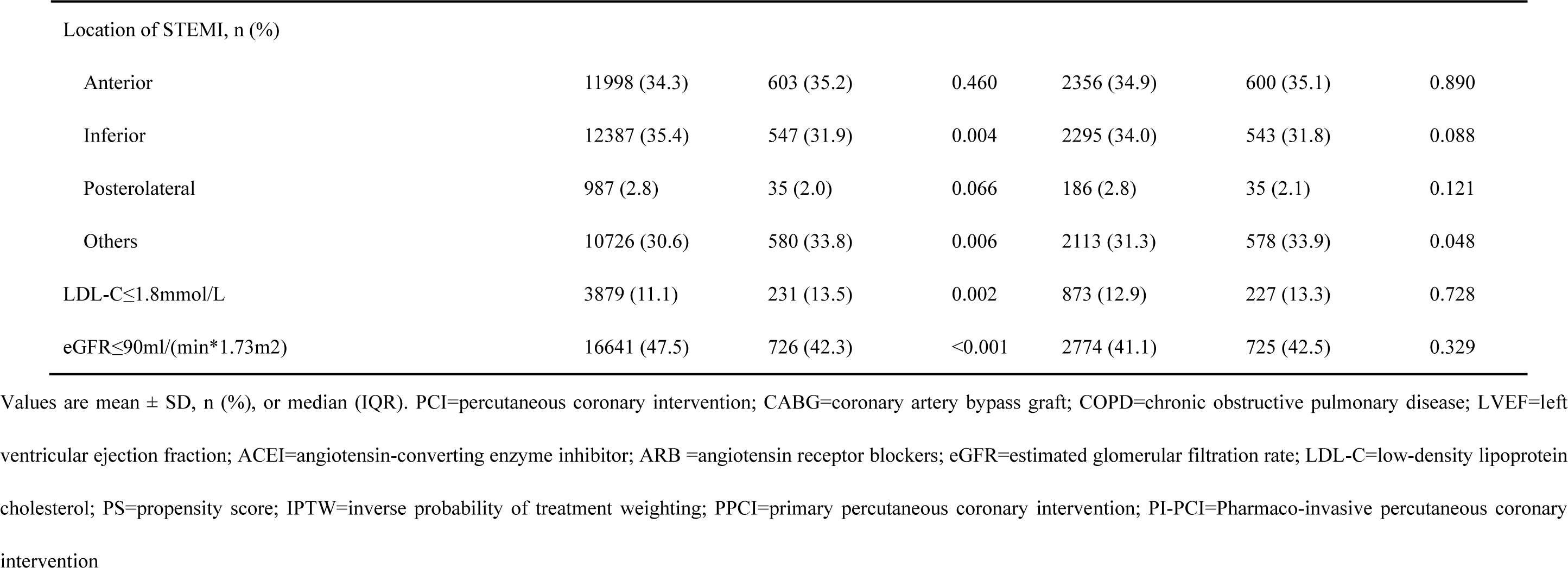
Comparison of baseline characteristics between PPCI and PI-PCI groups in the whole, PS-matched and IPTW cohorts.

**Supplemental Table 3.**
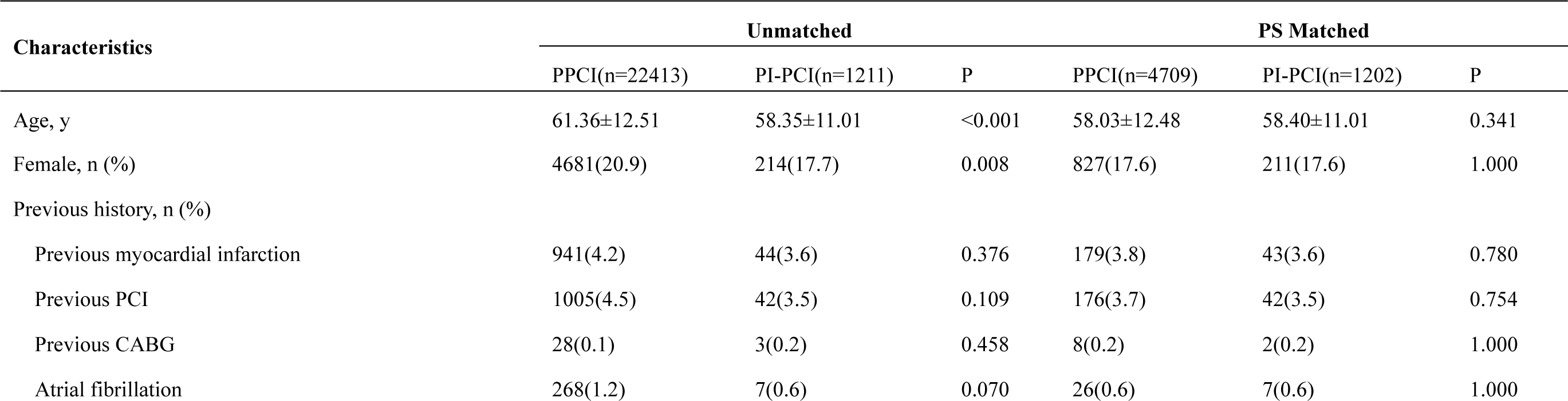
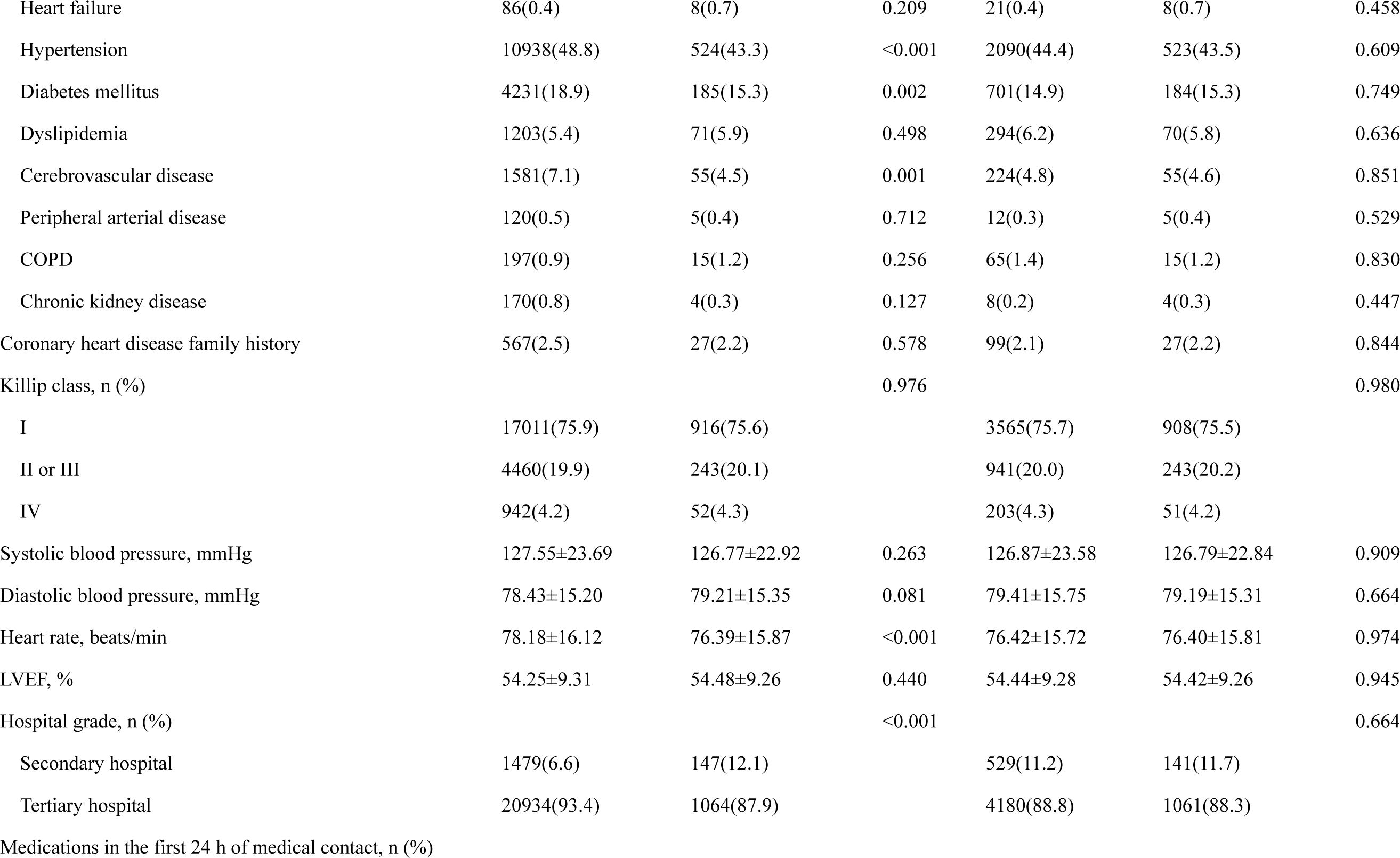
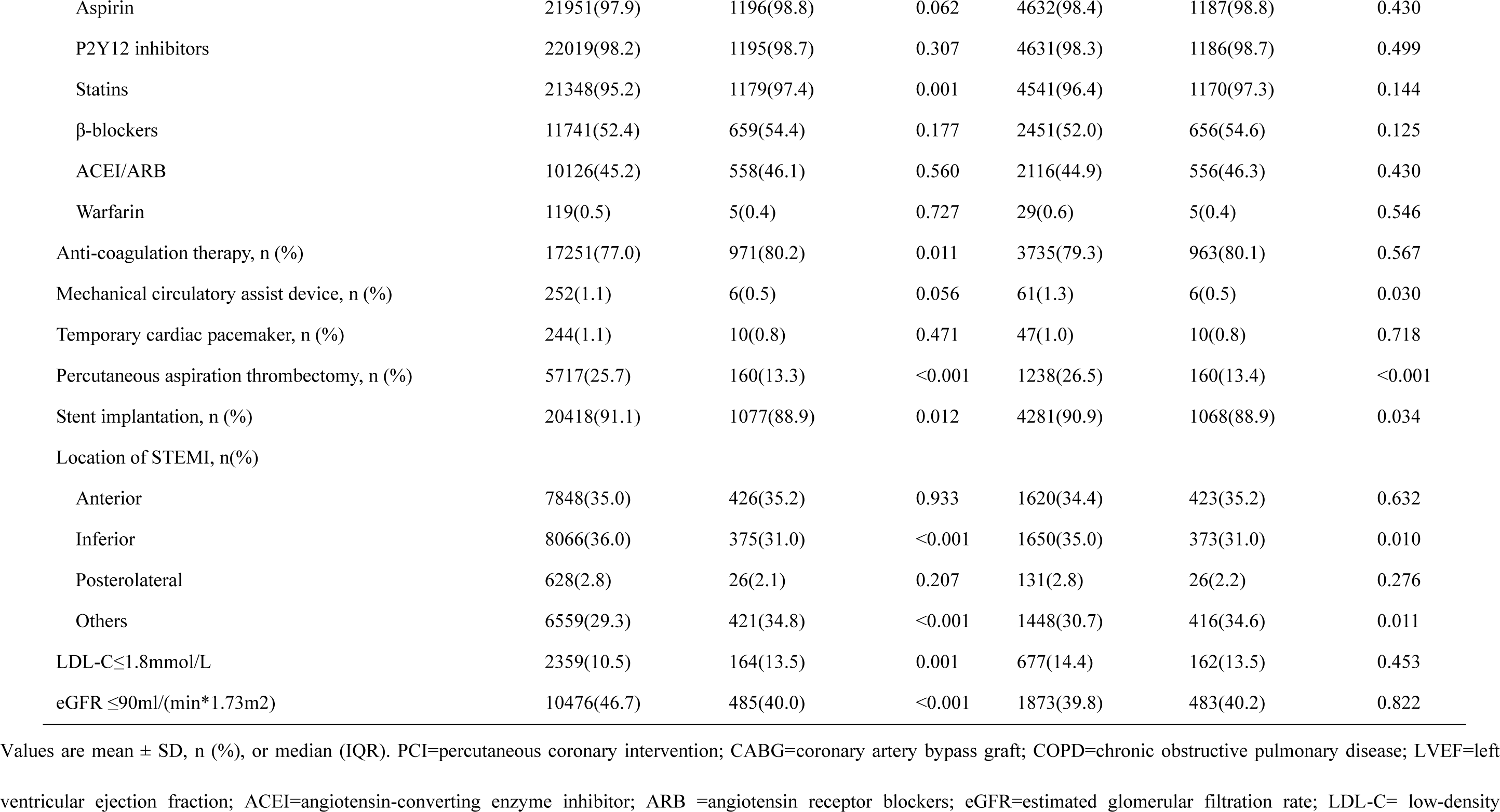
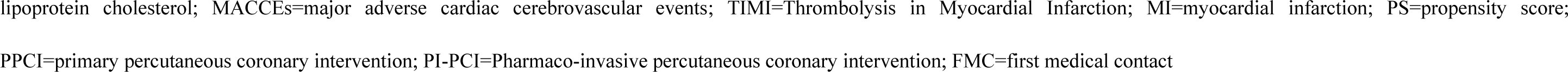
Comparison of baseline characteristics between PPCI group and PI-PCI group in the time from onset to FMC ≥ 3h.

**Supplemental Table 4.**
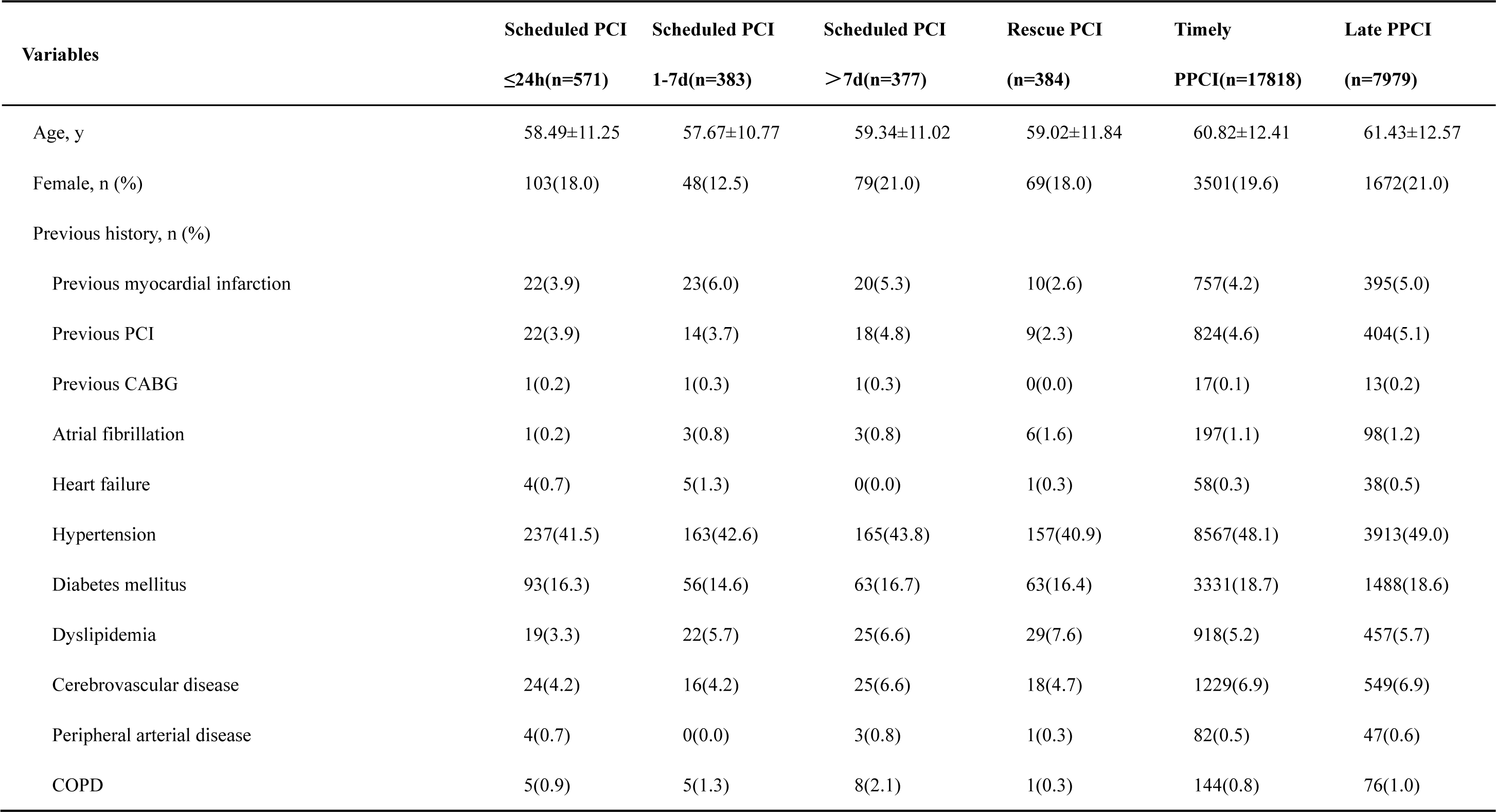
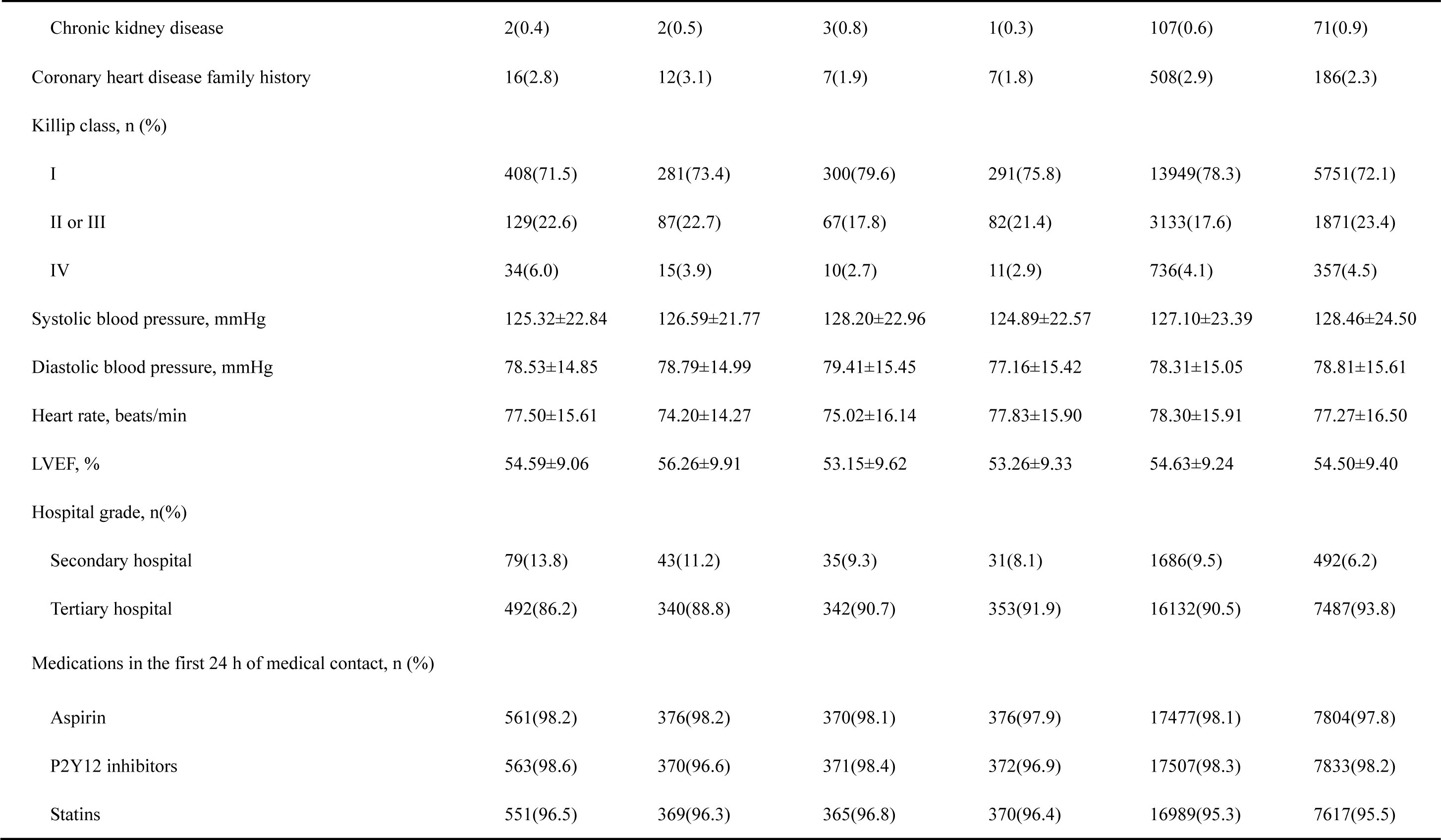
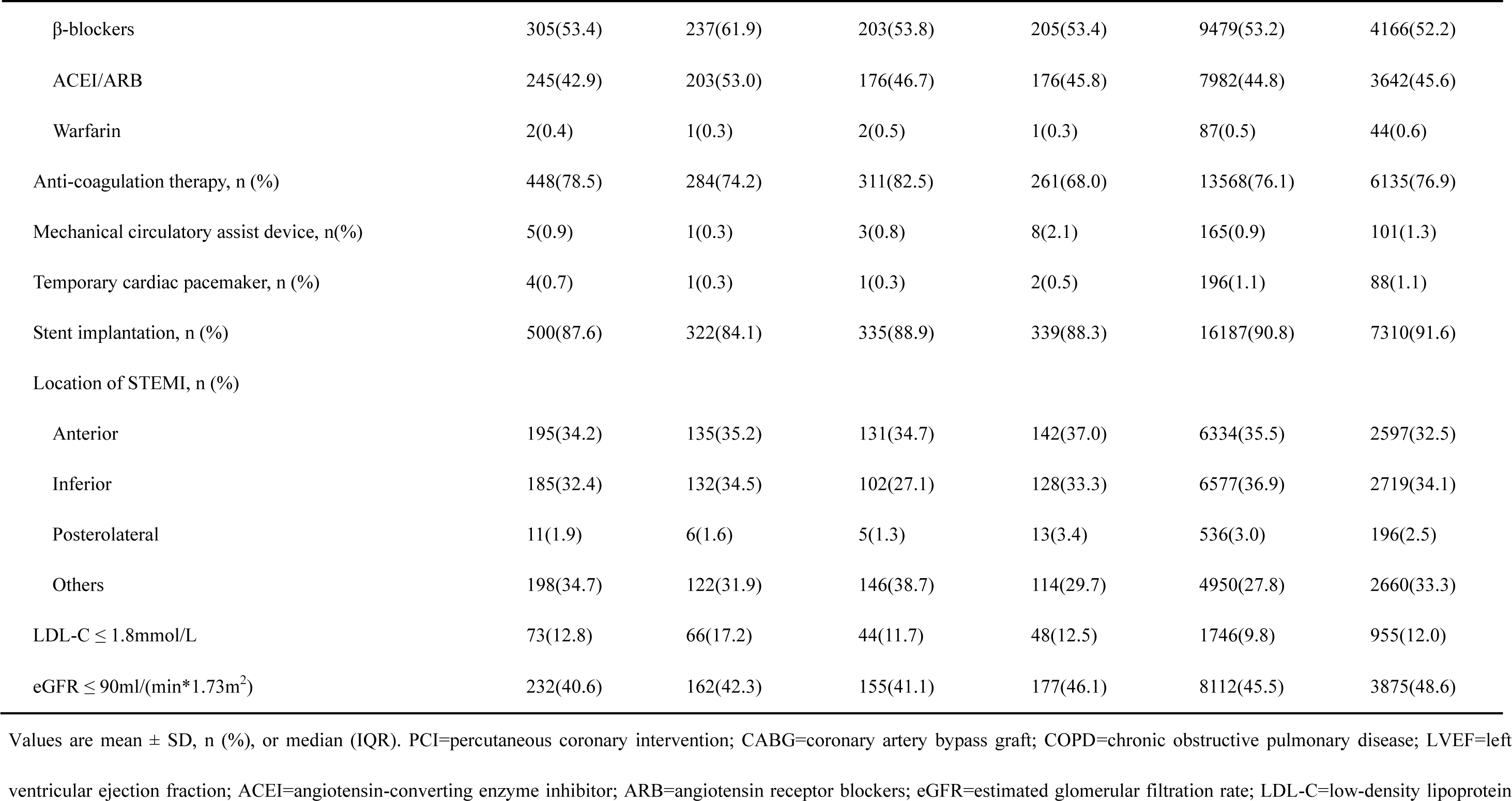
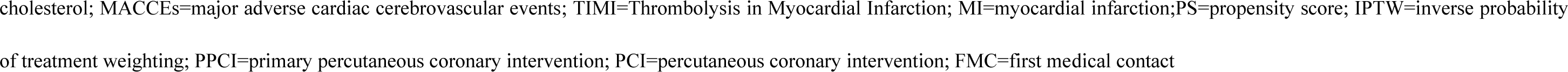
Comparison of baseline characteristics between subgroups.

**Supplemental Table 5.**
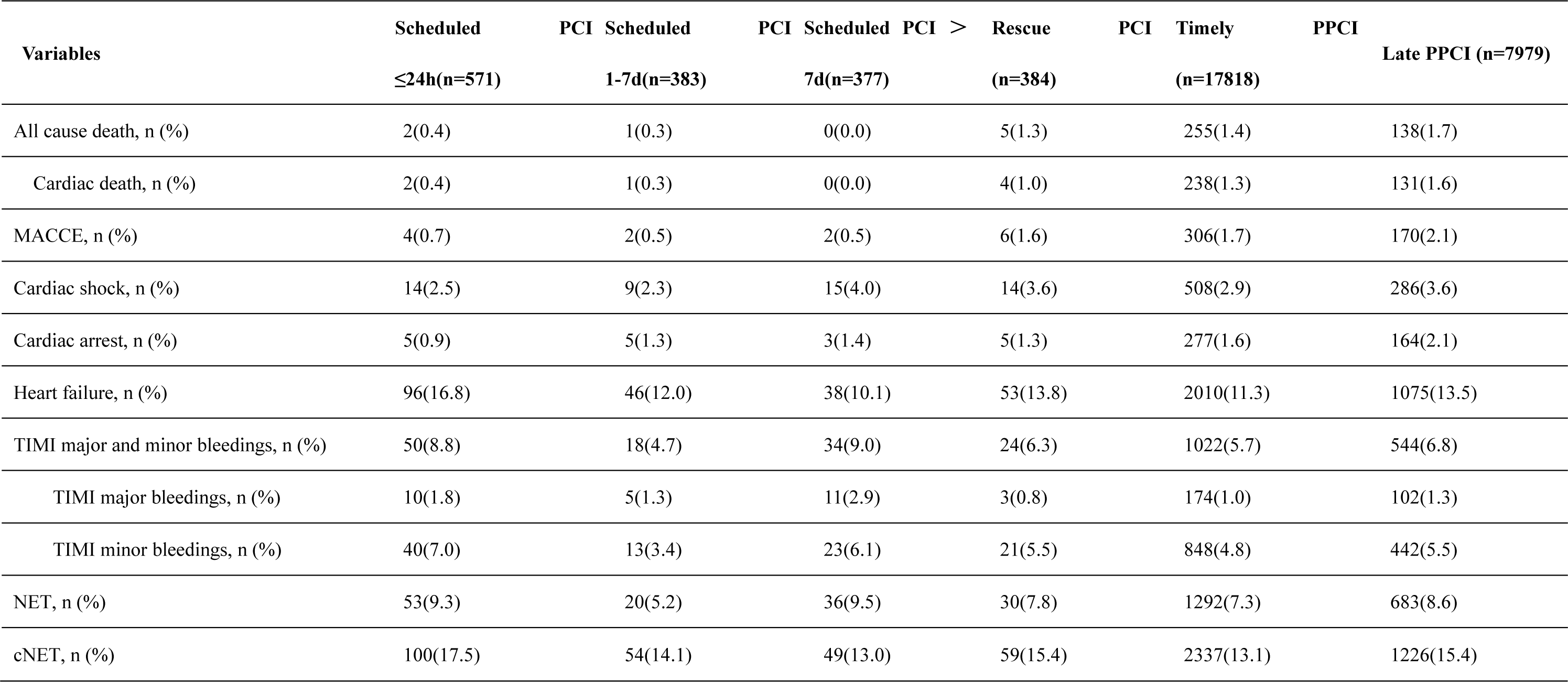
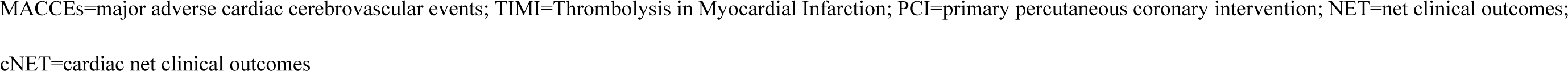
In-hospital outcomes between subgroups.

**Supplemental Table 6.** All participating hospitals.

| **ID** | **Hospitals** | **Territories** | **Provinces** | **City** | **Investigator** |
| --- | --- | --- | --- | --- | --- |
| 1 | Yangzhou First People's Hospital | Eastern China | Jiangsu | Yangzhou | Aihua Li |
| 2 | Shanxi Cardiovascular Hospital | Northern China | Shanxi | Taiyuan | Bao Li |
| 3 | Nanjing Drum Tower Hospital, The Affiliated Hospital of Nanjing University Medical School | Eastern China | Jiangsu | Nanjing | Biao Xu, Guangshu Han |
| 4 | Hainan General Hospital | Southern China | Hainan | Haikou | Bin Li |
| 5 | The Second Hospital of Jilin University | Northeast China | Jilin | Changchun | Bin Liu |
| 6 | Shanghai Jingan District Shibei Hospital | Eastern China | Shanghai | Shanghai | Bin Wang |
| 7 | Guangyuan Central Hospital | Northwest China | Sichuan | Guangyuan | Bing Fu |
| 8 | The Second Affiliated Hospital of Harbin Medical University | Northeast China | Heilongjiang | Harbin | Bo Yu |
| 9 | The 463 Hospital of Chinese People's Liberation Army | Northeast China | Liaoning | Shenyang | Bosong Yang |
| 10 | The Mianyang Central Hospital | Northwest China | Sichuan | Mianyang | Caidong Luo |
| 11 | The Ninth Hospital Affiliated to Shanghai Jiaotong University School of Medicine | Eastern China | Shanghai | Shanghai | Changqian Wang |
| 12 | Zhangzhou Municipal Hospital of Fujian Province | Eastern China | Fujian | Zhangzhou | Changyong Liu |
| 13 | Shimen People's Hospital | Central China | Hunan | Changde | Chuanliang Liang |
| 14 | Henan Provincial People's Hospital | Central China | Henan | Zhengzhou | Chuanyu Gao |
| 15 | Shanxi Provincial People's Hospital | Northern China | Shanxi | Taiyuan | Chunlin Lai |
| 16 | Xihua County People's Hospital | Central China | Henan | Zhoukou | Chuntong Wang |
| 17 | Liaocheng People's Hospital | Eastern China | Shandong | Liaocheng | Chunyan Zhang |
| 18 | The Third People's Hospital of Yancheng City | Eastern China | Jiangsu | Yancheng | Chunyang Wu |
| 19 | Quyang Renji Hospital | Northern China | Hebei | Baoding | Congliang Zhang |
| 20 | Xinqiao Hospital, Third Military Medical University | Southwest China | Chongqing | Chongqing | Cui Bin, Lan Huang |
| 21 | The Second Xiangya Hospital of Central South University | Central China | Hunan | Changsha | Daoquan Peng |
| 22 | The Central Hospital of Panzhihua | Northwest China | Sichuan | Panzhihua | Dawen Xu |
| 23 | China Meitan General Hospital | Northern China | Beijing | Beijing | Di Wu |
| 24 | Xiantao First People's Hospital | Central China | Hubei | Xiantao | Dongmei Zhu |
| 25 | Chest Hospital of Xinjiang Uygur Autonomous Region | Northwest China | Xinjiang | Urumchi | Dongsheng Chai |
| 26 | Beian First People's Hospital | Northeast China | Heilongjiang | Heihe | Dongyan Li |
| 27 | The 309th Hospital of Chinese People's Liberation Army | Northern China | Beijing | Beijing | Fakuan Tang, Jun Xiao |
| 28 | Baiyin City Center Hospital | Northwest China | Gansu | Baiyin | Fang Zhao |
| 29 | Deqing People's Hospital | Eastern China | Zhejiang | Huzhou | Fangfang Huang |
| 30 | Dunhua City Hospital | Northeast China | Jilin | Yanbian | Fanju Meng |
| 31 | Suizhou Central Hospital | Central China | Hubei | Suizhou | Fengwei Li |
| 32 | Binyang People's Hospital | Southern China | Guangxi | Nanning | Fudong Gan |
| 33 | The First Hospital of Qiqihaer City | Northeast China | Heilongjiang | Qiqihaer | Gang Xu |
| 34 | The Third People's Hospital of Bengbu City | Eastern China | Anhui | Bengbu | Gengsheng Sang |
| 35 | Zhongda Hospital, Southeast University | Eastern China | Jiangsu | Nanjing | Genshan Ma |
| 36 | The First Hospital of Jiamusi City | Northeast China | Heilongjiang | Jiamusi | Guixia Zhang |
| 37 | The First Affiliated Hospital of Liaoning Medical University | Northeast China | Liaoning | Jinzhou | Guizhou Tao |
| 38 | Luan County People's Hospital | Northern China | Hebei | Tangshan | Guo Li |
| 39 | Guiding People's Hospital | Southwest China | Guizhou | Qinan | Guoduo Chen |
| 40 | Haidong Ping'an District Hospital of Traditional Chinese Medicine | Northwest China | Qinghai | Haidong | Guoqin Xin |
| 41 | Xinjiang Uygur Autonomous Region People's Hospital | Northwest China | Xinjiang | Urumchi | Guoqing Li |
| 42 | Sir Run Run Shaw Hospital, College of Medicine, Zhejiang University | Eastern China | Zhejiang | Hangzhou | Guosheng Fu |
| 43 | Zhoushan People's Hospital | Eastern China | Zhejiang | Zhoushan | Guoxiong Chen |
| 44 | Dalian Municipal Central Hospital | Northeast China | Liaoning | Dalian | Hailong Lin |
| 45 | Hebei Daming County People's Hospital | Northern China | Hebei | Handan | Haiping Guo |
| 46 | Dongguan Changping hospital | Southern China | Guangdong | Dongguan | Haiyun Lin |
| 47 | Renmin Hospital of Wuhan University | Central China | Hubei | Wuhan | Hong Jiang |
| 48 | Honghu People's Hospital | Central China | Hubei | Jingzhou | Hong Liu |
| 49 | Ningxia People's Hospital | Northwest China | Ningxia | Yinchuan | Hong Luan |
| 50 | The First People's Hospital of Yunnan Province (Kunhua Hospital) | Northwest China | Yunnan | Kunming | Hong Zhang |
| 51 | The People's Hospital Feixian | Eastern China | Shandong | Linyi | Honghua Deng |
| 52 | Beijing Friendship Hospital, Capital Medical University | Northern China | Beijing | Beijing | Hongwei Li |
| 53 | The First Affiliated Hospital of Bengbu Medical College | Eastern China | Anhui | Bengbu | Honhju Wang |
| 54 | The Central Hospital of Zhoukou | Central China | Henan | Zhoukou | Hualing Liu |
| 55 | Nanpi People's Hospital | Northern China | Hebei | Cangzhou | Hui Dong |
| 56 | Anyang District Hospital | Central China | Henan | Anyang | Hui Liu |
| 57 | Dalian Fourth People's Hospital | Northeast China | Liaoning | Dalian | Huifang Zhang |
| 58 | General Hospital of TISCO | Northern China | Shanxi | Taiyuan | Huifeng Wang |
| 59 | Ningbo First Hospital | Eastern China | Zhejiang | Ningbo | Huimin Chu |
| 60 | Huining People's Hospital | Northwest China | Gansu | Baiyin | Jiabin Xi |
| 61 | Jining City Yanzhou District People's Hospital | Eastern China | Shandong | Jining | Jian Yang |
| 62 | Dongguan People's Hospital | Southern China | Guangdong | Dongguan | Jianfeng Ye |
| 63 | Panyu Hospital of Chinese Medicine | Southern China | Guangdong | Guangzhou | Jianhao Li |
| 64 | Sichuan Provincial People's Hospital | Northwest China | Sichuan | Chengdu | Jianhong Tao |
| 65 | Mudanjiang Cardiovascular Disease Hospital | Northeast China | Heilongjiang | Mudanjiang | Jianwen Liu |
| 66 | People's Hospital of Wugang | Central China | Hunan | Shaoyang | JiaoMei Yang |
| 67 | Yichang Central Hospital | Central China | Hubei | Yichang | Jiawang Ding |
| 68 | Zhongda Hospital, Southeast University (Jiangbei) | Eastern China | Jiangsu | Nanjing | Jiayi Tong |
| 69 | People's Hospital of Rongchang District | Southwest China | Chongqing | Chongqing | Jie Chen |
| 70 | The First Hospital of Peking University | Northern China | Beijing | Beijing | Jie Jiang |
| 71 | Ye County people's hospital | Central China | Henan | Pingdingshan | Jie Yang |
| 72 | Qilu Hospital of Shandong University | Eastern China | Shandong | Jinan | Jifu Li |
| 73 | The Affiliated Hospital of Jiangsu University | Eastern China | Jiangsu | Zhenjiang | Jinchuan Yan |
| 74 | Wuhan University of Science and Technology Hospital | Central China | Hubei | Wuhan | Jing Hu |
| 75 | Shenyang City Electricity Central Hospital | Northeast China | Liaoning | Shenyang | Jing Xu |
| 76 | Sun Yat-sen Memorial Hospital, Sun Yat-sen University | Southern China | Guangdong | Guangzhou | Jingfeng Wang |
| 77 | Yuncheng Hospital | Eastern China | Shandong | Heze | Jinglan Diao |
| 78 | Fengrun District Second People's Hospital | Northern China | Hebei | Tangshan | Jingshan Zhao |
| 79 | The First People's Hospital of Nanning City | Southern China | Guangxi | Nanning | Jinru Wei |
| 80 | Zhangping City Hospital | Eastern China | Fujian | Longyan | Jinxing Yi |
| 81 | The First Affiliated Hospital of Fujian Medical University | Eastern China | Fujian | Fuzhou | Jinzi Su |
| 82 | Chengdu Third People's Hospital | Northwest China | Sichuan | Chengdu | Jiong Tang |
| 83 | Guangdong General Hospital | Southern China | Guangdong | Guangzhou | Jiyan Chen |
| 84 | Fujin City Central Hospital of Heilongjiang Province | Northeast China | Heilongjiang | Jiamusi | Jiyan Yin |
| 85 | Yantaishan hospital | Eastern China | Shandong | Yantai | Juexin Fan |
| 86 | Qingdao Municipal Hospital | Eastern China | Shandong | Qingdao | Jun Guan |
| 87 | Zhongshan Hospital Affiliated to Fudan University | Eastern China | Shanghai | Shanghai | Junbo Ge |
| 88 | Hospital of Xinjiang Production & Construction Corps | Northwest China | Xinjiang | Urumchi | Junming Liu |
| 89 | Linfen People's Hospital | Northern China | Shanxi | Linfen | Junping Deng |
| 90 | The First People's Hospital of Horqin District, Tongliao City | Northern China | Inner Mongolia | Tongliao | Junping Fang |
| 91 | The Military General Hospital of Beijing PLA | Northern China | Beijing | Beijing | Junxia Li |
| 92 | Longyan First Hospital | Eastern China | Fujian | Longyan | Kaihong Chen |
| 93 | Guiyang Sixth People's Hospital | Southwest China | Guizhou | Guiyang | Kalan Luo |
| 94 | Affiliated Hospital of Guangdong Medical College | Southern China | Guangdong | Guangzhou | Keng Wu |
| 95 | Jiangxi Provincial People's Hospital | Eastern China | Jiangxi | Nanchang | Lang Ji |
| 96 | The First Affiliated Hospital of Guangxi Medical University | Southern China | Guangxi | Nanning | Lang Li |
| 97 | Tongren Hospital Affiliated to Shanghai Jiaotong University School of Medicine | Eastern China | Shanghai | Shanghai | Li Jiang |
| 98 | Huaiyang People's Hospital | Central China | Henan | Zhoukou | Li Wei |
| 99 | Binzhou City Center Hospital | Eastern China | Shandong | Binzhou | Lijun Meng |
| 100 | Anhui Provincial Hospital | Eastern China | Anhui | Hefei | Likun Ma |
| 101 | Xiangtan City Central Hospital | Central China | Hunan | Xiangtan | Lilong Tang |
| 102 | Tangshan City Fengrun District People's Hospital | Northern China | Hebei | Tangshan | Lin Wang |
| 103 | The First Hospital of Haerbin City | Northeast China | Heilongjiang | Harbin | Lin Wei |
| 104 | The First Affiliated Hospital of Zhengzhou University | Central China | Henan | Zhengzhou | Ling Li |
| 105 | Xijing Hospital | Northwest China | Shaanxi | Xi'an | Ling Tao |
| 106 | Yiniang Hospital | Southwest China | Yunnan | Kunming | Liqiong Yang |
| 107 | The Affiliated Hospital of Guizhou Medical University | Southwest China | Guizhou | Guiyang | Lirong Wu |
| 108 | Central Hospital Affiliated to Shenyang Medical College | Northeast China | Liaoning | Shenyang | ManZhang,<br>Kaiming Chen |
| 109 | Hepu People's Hospital | Southern China | Guangxi | Beihai | Meisheng Lai |
| 110 | First Affiliated Hospital of the People's Liberation Army General Hospital | Northern China | Beijing | Beijing | Miao Tian |
| 111 | Yanting People's Hospital | Southwest China | Sichuan | Mianyang | Mingcheng Bai |
| 112 | The Second People's Hospital of Yunnan Province | Southwest China | Yunnan | Kunming | Minghua Han |
| 113 | Haikou People's Hospital | Southern China | Hainan | Haikou | Moshui Chen |
| 114 | Geological Mining Hospital of Hunan Province | Central China | Hunan | Changsha | Naiyi Liang |
| 115 | The Eight Affiliated Hospital, Sun Yat-sen University | Southern China | Guangdong | Guangzhou | Nan Jia |
| 116 | The Central Hospital of Xuzhou | Eastern China | Jiangsu | Xuzhou | Peiying Zhang |
| 117 | The Second hospital of Dalian Medical University | Northeast China | Liaoning | Dalian | Peng Qu |
| 118 | The second people's hospital of Mengcheng | Eastern China | Anhui | Bozhou | Pengfei Zhang |
| 119 | Fuqing Cite Hospital | Eastern China | Fujian | Fuqing | Ping Chen |
| 120 | The First Affiliated Hospital of Liaoning University of Traditional Chinese Medicine | Northeast China | Liaoning | Shenyang | Ping Hou |
| 121 | Gansu Provincial Hospital | Northwest China | Gansu | Lanzhou | Ping Xie |
| 122 | Beijing Tsinghua Changgung Hospital | Northern China | Beijing | Beijing | Ping Zhang |
| 123 | The First Affiliated Hospital of Henan University of Science and Technology | Central China | Henan | Luoyang | Pingshuan Dong |
| 124 | Guizhou Provincial People's Hospital | Northwest China | Guizhou | Guiyang | Qiang Wu |
| 125 | The First Affiliated Hospital of Xiamen University | Eastern China | Fujian | Xiamen | Qiang Xie |
| 126 | Chenzhou First People's Hospital | Central China | Hunan | Chenzhou | Qiaoqing Zhong |
| 127 | Lujiang People's Hospital | Eastern China | Anhui | Hefei | Qichun Wang |
| 128 | Yuzhou City Central Hospital | Central China | Henan | Xuchang | Qinfeng Su |
| 129 | People's Hospital of Qinghai Province | Northwest China | Qinghai | Xining | Rong Chang |
| 130 | Quanzhou First Hospital | Eastern China | Fujian | Quanzhou | Rong Lin |
| 131 | Baotou City Center Hospital | Northern China | Inner Mongolia | Baotou | Ruiping Zhao |
| 132 | Affiliated Hospital of Ningxia Medical University | Northwest China | Ningxia | Yinchuan | Shaobin Jia |
| 133 | Beijing Anzhen Hospital, Capital Medical University | Northern China | Beijing | Beijing | Shaoping Nie |
| 134 | Wuzhou People's Hospital | Southern China | Guangxi | Wuzhou | Shaowu Ye |
| 135 | North Jiangsu People's Hospital | Eastern China | Jiangsu | Yangzhou | Shenghu He |
| 136 | People's Hospital of Bozhou District | Southwest China | Guizhou | Zunyi | Shengyong Chen |
| 137 | Shanghai Sixth People's Hospital | Eastern China | Shanghai | Shanghai | Shixin Ma |
| 138 | The Central Hospital of Jilin | Northeast China | Jilin | Changchun | Shuangbin Li |
| 139 | The First Hospital of Handan | Northern China | Hebei | Handan | Shuanli Xin |
| 140 | The Fourth Affiliated Hospital Zhejiang University School of Medicine | Eastern China | Zhejiang | Yiwu | Shudong Xia |
| 141 | Nenjiang People's Hospital | Northeast China | Heilongjiang | Heihe | Shuhua Zhang |
| 142 | Duzishan Petrochemical Hospital | Northwest China | Xinjiang | Karamay | Shuqiu Qu |
| 143 | Huai'an First People's Hospital | Eastern China | Jiangsu | Huai'an | Shuren Ma |
| 144 | Hunan Changsha County First People's Hospital | Central China | Hunan | Changsha | Siding Wang |
| 145 | Li County Hospital of Traditional Chinese Medicine | Central China | Hunan | Changde | Songbai Li |
| 146 | The First Affiliated Hospital of Chongqing Medical University | Southwest China | Chongqing | Chongqing | Suxin Luo |
| 147 | Nanchong Central Hospital | Northwest China | Sichuan | Nanchong | Tao Liu |
| 148 | Ningjin People's Hospital | Eastern China | Shandong | Dezhou | Tao Zhang |
| 149 | Guang'an People's Hospital | Southwest China | Sichuan | Guang'an | Tian Tuo |
| 150 | Navy General Hospital | Northern China | Beijing | Beijing | Tianchang Li |
| 151 | Xiangya Hospital Central South University | Central China | Hunan | Changsha | Tianlun Yang |
| 152 | Gongyi people's hospital | Central China | Henan | Zhengzhou | Tianmin Du |
| 153 | Guangzhou Red Cross Hospital | Southern China | Guangdong | Guangzhou | Tongguo Wu |
| 154 | Dongfeng Hospital | Northeast China | Jilin | Liaoyuan | Wei Liu |
| 155 | Zhejiang Provincial Hospital of TCM | Eastern China | Zhejiang | Hangzhou | Wei Mao |
| 156 | The First People's Hospital of Longquanyi District | Southwest China | Sichuan | Chengdu | Wei Tuo |
| 157 | The First Affiliated Hospital of Guangzhou Medical College | Southern China | Guangdong | Guangzhou | Wei Wang |
| 158 | The Third Xiangya Hospital of Central South University | Central China | Hunan | Changsha | Weihong Jiang |
| 159 | The First Affiliated Hospital of Wenzhou Medical University | Eastern China | Zhejiang | Wenzhou | Weijian Huang |
| 160 | Affiliated Hospital of Qinghai University | Northwest China | Qinghai | Xining | Weijun Liu |
| 161 | Jianshui County People's Hospital | Southwest China | Yunnan | Honghe | Weiqing Fan |
| 162 | The Second Affiliated Hospital of Soochow University | Eastern China | Jiangsu | Suzhou | Weiting Xu |
| 163 | Teda International Cardiovascular Hospital | Northern China | Tianjin | Tianjin | Wenhua Lin |
| 164 | Wuhan Asia Heart Hospital | Central China | Hubei | Wuhan | Xi Su |
| 165 | Shanghai Jiading District Center Hospital | Eastern China | Shanghai | Shanghai | Xia Chen |
| 166 | Guangxi Hengxian County People's Hospital | Southern China | Guangxi | Nanning | Xianan Zhang |
| 167 | The Second Hospital of Hebei Medical University | Northern China | Hebei | Shijiazhuang | Xianghua Fu |
| 168 | The First Affiliated Hospital of Soochow University | Eastern China | Jiangsu | Suzhou | Xiangjun Yang |
| 169 | Changhai Hospital of Shanghai | Eastern China | Shanghai | Shanghai | Xianxian Zhao |
| 170 | Affiliated Hospital of Yan'an University | Northwest China | Shaanxi | Yan'an | Xiaochuan Ma |
| 171 | The First People's Hospital of Jining | Eastern China | Shandong | Jining | Xiaofei Sun |
| 172 | Longhui County People's Hospital | Central China | Hunan | Shaoyang | Xiaojun Wang |
| 173 | Tonglu First People's Hospital | Eastern China | Zhejiang | Hangzhou | Xiaolan Li |
| 174 | Xinmi people's hospital | Central China | Henan | Zhengzhou | Xiaolei Li |
| 175 | Zunhua People's Hospital | Northern China | Hebei | Tangshan | Xiaoli Yang |
| 176 | West China Hospital of Sichuan University | Northwest China | Sichuan | Chengdu | Xiaoping Chen |
| 177 | The Central Hospital of Taiyuan | Northern China | Shanxi | Taiyuan | Xiaoping Chen |
| 178 | Datong City Second People's Hospital | Northern China | Shanxi | Datong | Xiaoqin Zhang |
| 179 | The Second Affiliated Hospital to Nanchang University | Eastern China | Jiangxi | Nanchang | Xiaoshu Cheng |
| 180 | Yuzhong County People's Hospital | Northwest China | Gansu | Lanzhou | Xiaowei Peng |
| 181 | Qinyang People's Hospital | Central China | Henan | Jiaozuo | Xiaowen Ma |
| 182 | Hebei General Hospital | Northern China | Hebei | Shijiazhuang | Xiaoyong Qi |
| 183 | Yutian Hospital | Northern China | Hebei | Tangshan | Xiaoyun Feng |
| 184 | The Third Affiliated Hospital of Guangzhou Medical College | Southern China | Guangdong | Guangzhou | Ximing Chen |
| 185 | Chongqing Hechuan District People's Hospital | Southwest China | Chongqing | Chongqing | Xin Tang |
| 186 | The First Affiliated Hospital of Wannan Medical College | Eastern China | Anhui | Wuhu | Xingsheng Tang |
| 187 | Inner Mongolia People's Hospital | Northern China | Inner Mongolia | Hohhot | Xingsheng Zhao |
| 188 | Ledong Second People's Hospital | Southern China | Hainan | Ledong | Xiufeng Chen |
| 189 | Wuxi Xishan People's Hospital | Eastern China | Jiangsu | Wuxi | Xudong Li |
| 190 | Tangdu Hospital of The Fourth Military Medical University | Northwest China | Shaanxi | Xi'an | Xue Li |
| 191 | Shanghai East Hospital Affiliated to Tongji University | Eastern China | Shanghai | Shanghai | Xuebo Liu |
| 192 | Beijing Fangshan District First Hospital | Northern China | Beijing | Beijing | Xuemei Peng |
| 193 | The General Hospital of Shenyang Military Region | Northeast China | Liaoning | Shenyang | Yaling Han |
| 194 | Xiamen Cardiovascular Disease Hospital | Eastern China | Fujian | Xiamen | Yan Wang |
| 195 | Tieli People's Hospital | Northeast China | Heilongjiang | Yichun | Yanbo Niu |
| 196 | Dianjiang People's Hospital | Southwest China | Chongqing | Chongqing | Yang Yu |
| 197 | The First Hospital of Jilin University | Northeast China | Jilin | Changchun | Yang Zheng |
| 198 | The Second Affiliated Hospital of Qiqihar Medical Hospital | Northeast China | Heilongjiang | Qiqihar | Yanli Wang |
| 199 | General Hospital of Guangzhou Military Command | Southern China | Guangdong | Guangzhou | Yanlie Zheng |
| 200 | Fujian Provincial Hospital | Eastern China | Fujian | Fuzhou | Yansong Guo |
| 201 | The First Affiliated hospital of Dalian Medical University | Northeast China | Liaoning | Dalian | Yanzong Yang |
| 202 | The First People's Hospital of Changde | Central China | Hunan | Changde | Yi Huang |
| 203 | Tianjin Chest Hospital | Northern China | Tianjin | Tianjin | Yin Liu |
| 204 | Hunan Provincial People's Hospital | Central China | Hunan | Changsha | Ying Guo |
| 205 | Longmen People's Hospital | Southern China | Guangdong | Huizhou | Yingchao Luo |
| 206 | People's Hospital of Yuxi City | Southwest China | Yunnan | Yuxi | Yinglu Hao |
| 207 | The First Affiliated Hospital of China Medical University | Northeast China | Liaoning | Shenyang | Yingxian Sun |
| 208 | The People's Hospital of Guangxi Zhuang Autonomous Region | Southern China | Guangxi | Nanning | Yingzhong Lin |
| 209 | The First Teaching Hospital of Xinjiang Medical University | Northwest China | Xinjiang | Urumchi | Yitong Ma |
| 210 | Dazhou Central Hospital | Northwest China | Sichuan | Dazhou | Yong Guo |
| 211 | Mingguang People's Hospital | Eastern China | Anhui | Chuzhou | Yong Li |
| 212 | Baogang Hospital | Northern China | Inner Mongolia | Baotou | Yongdong Li |
| 213 | Jiangsu Binhai County People's Hospital | Eastern China | jiangsu | Yancheng | Yonglin Zhang |
| 214 | The Fourth Affiliated Hospital of China Medical University | Northeast China | Liaoning | Shenyang | Yuanzhe Jin |
| 215 | First Affiliated Hospital of Harbin Medical University. | Northeast China | Heilongjiang | Harbin | Yue Li |
| 216 | Sihui People's Hospital | Southern China | Guangdong | Zhaoqing | Yuehua Huang |
| 217 | Tianjin Medical University General Hospital | Northern China | Tianjin | Tianjin | Yuemin Sun |
| 218 | Qian'an People's Hospital | Northern China | Hebei | Tangshan | Yuheng Yang |
| 219 | Zhalantun People's Hospital | Northern China | Inner Mongolia | Hulunbeier | Yuhua Zhu |
| 220 | Longjiang First People's Hospital | Northeast China | Heilongjiang | Qiqihar | Yuhuan Shi |
| 221 | The Second Affiliated Hospital of Zhengzhou University | Central China | Henan | Zhengzhou | Yulan Zhao |
| 222 | Nanfang Hospital of Southern Medical University | Southern China | Guangdong | Guangzhou | Yuqing Hou |
| 223 | The First Affiliated Hospital to Nanchang University | Eastern China | Jiangxi | Nanchang | Zeqi Zheng |
| 224 | Cangzhou Central Hospital | Northern China | Hebei | Cangzhou | Zesheng Xu |
| 225 | The Central Hospital of Shaoyang | Central China | Hunan | Shaoyang | Zewei Ouyang |
| 226 | Yulong Hospital | Southwest China | Yunnan | Lijiang | Zeyuan He |
| 227 | Affiliated Hospital of North Sichuan Medical College | Northwest China | Sichuan | Nanchong | Zhan Lv |
| 228 | The People's Hospital of Liaoning Province | Northeast China | Liaoning | Shenyang | Zhanquan Li |
| 229 | The First Affiliated Hospital of Jiamusi University | Northeast China | Heilongjiang | Jiamusi | Zhaofa He |
| 230 | Tangshan Gongren Hospital | Northern China | Hebei | Tangshan | Zheng Ji |
| 231 | The First Affiliated Hospital of Lanzhou University | Northwest China | Gansu | Lanzhou | Zheng Zhang |
| 232 | The Third Hospital of Shijiazhuang | Northern China | Hebei | Shijiazhuang | Zhenguo Ji |
| 233 | Huaibei Miners General Hospital | Eastern China | Anhui | Huaibei | Zhenqi Su |
| 234 | Wuxi People's Hospital | Eastern China | Jiangsu | Wuxi | Zhenyu Yang |
| 235 | Linyi People's Hospital | Eastern China | Shandong | Linyi | Zhihong Ou |
| 236 | Jiangsu Province Hospital | Eastern China | Jiangsu | Nanjing | Zhijian Yang |
| 237 | The Second Hospital of Shanxi Medical University | Northern China | Shanxi | Taiyuan | Zhiming Yang |
| 238 | The Affiliated Hospital of Xuzhou Medical College | Eastern China | Jiangsu | Xuzhou | Zhirong Wang |
| 239 | Southwest Hospital, Third Military Medical University | Southwest China | Chongqing | Chongqing | Zhiyuan Song |
| 240 | Zhijin People's Hospital | Southwest China | Guizhou | Bijie | Zhongshan Wang |
| 241 | The First Affiliated Hospital of Xi'an Jiaotong University | Northwest China | Shaanxi | Xi'an | Zuyi Yuan |

## References

1. Thygesen K, Alpert JS, Jaffe AS, et al; Executive Group on behalf of the Joint European Society of Cardiology (ESC)/American College of Cardiology (ACC)/American Heart Association (AHA)/World Heart Federation (WHF) Task Force for the Universal Definition of Myocardial Infarction. Fourth Universal Definition of Myocardial Infarction (2018). Circulation 2018;138(20):e618–e651

2. The National Center for Cardiovascular Diseases. Interpretation of Report on Cardiovascular Health and Disease in China 2021. Chinese Circulation Journal, 2022, 36: 553-578. DOI:10.3969/j.issn.1000-3614.2022.06.001.

3. Gershlick AH, Banning AP, Myat A. Reperfusion therapy for STEMI: is there still a role for thrombolysis in the era of primary percutaneous coronary intervention? Lancet. 2013;382:624–632. doi: 10.1016/S0140-6736(13)61454-3.

4. Steg PG, James SK, Atar D, Badano LP, Blomstrom-Lundqvist C, Borger MA, Di Mario C, Dickstein K, Ducrocq G, Fernandez-Aviles F, Gershlick AH, Giannuzzi P, Halvorsen S, Huber K, Juni P, Kastrati A, Knuuti J, Lenzen MJ, Mahaffey KW, Valgimigli M, van ‘t Hof A, Widimsky P, Zahger D. ESC guidelines for the management of acute myocardial infarction in patients presenting with ST-segment elevation. Eur Heart J. 2012;33:2569–2619.

5. O’Gara PT, Kushner FG, Ascheim DD, Casey DE Jr, Chung MK, de Lemos JA, Ettinger SM, Fang JC, Fesmire FM, Franklin BA, Granger CB, Krumholz HM, Linderbaum JA, Morrow DA, Newby LK, Ornato JP, Ou N, Radford MJ, Tamis-Holland JE, Tommaso CL, Tracy CM, Woo YJ, Zhao DX, Anderson JL, Jacobs AK, Halperin JL, Albert NM, Brindis RG, Creager MA, DeMets D, Guyton RA, Hochman JS, Kovacs RJ, Kushner FG, Ohman EM, Stevenson WG, Yancy CW; American College of Cardiology Foundation/American Heart Association Task Force on Practice Guidelines. 2013 ACCF/AHA guideline for the management of ST-elevation myocardial infarction: a report of the American College of Cardiology Foundation/ American Heart Association Task Force on Practice Guidelines. Circulation. 2013;127:e362–e425. doi: 10.1161/CIR.0b013e3182742cf6.

6. Hao Y, Liu J, Liu J, et al. Rationale and design of the Improving Care for Cardiovascular Disease in China (CCC) project: a national effort to prompt quality enhancement for acute coronary syndrome. Am Heart J. 2016;179:107–115.

7. Chinese Society of Cardiology of Chinese Medical Association; Editorial Board of Chinese Journal of Cardiology. [2019 Chinese Society of Cardiology (CSC) guidelines for the diagnosis and management of patients with ST-segment elevation myocardial infarction]. Zhonghua Xin Xue Guan Bing Za Zhi. 2019 Oct 24;47(10):766-783. Chinese. doi: 10.3760/cma.j.issn.0253-3758.2019.10.003. PMID: 31648459.

8. 8. Chinese Society of Cardiology of Chinese Medical Association; Editorial Board of Chinese Journal of Cardiology. [Guideline and consensus for the management of patients with non-ST-elevation acute coronary syndrome(2016)]. Zhonghua Xin Xue Guan Bing Za Zhi. 2017 May 24;45(5):359-376. Chinese. doi:10.3760/cma.j.issn.0253-3758.2017.05.003. PMID: 28511320.

9. Chesebro JH, Knatterud G, Roberts R, et al. Thrombolysis in Myocardial Infarction (TIMI) Trial, Phase I: a comparison between intravenous tissue plasminogen activator and intravenous streptokinase. Clinical fifindings through hospital discharge. Circulation. 1987;76:142–154.

10. Xu H, Yang Y, Wang C, Yang J, Li W, Zhang X, Ye Y, Dong Q, Fu R, Sun H, Yan X, Gao X, Wang Y, Jia X, Sun Y, Wu Y, Zhang J, Zhao W, Sabatine MS, Wiviott SD; China Acute Myocardial Infarction Registry Investigators. Association of Hospital-Level Differences in Care With Outcomes Among Patients With Acute ST-Segment Elevation Myocardial Infarction in China. JAMA Netw Open. 2020 Oct 1;3(10):e2021677. doi: 10.1001/jamanetworkopen.2020.21677. PMID: 33095249; PMCID: PMC7584928.

11. Wu C, Li L, Wang S, Zeng J, Yang J, Xu H, Zhao Y, Wang Y, Li W, Jin C, Gao X, Yang Y, Qiao S. Fibrinolytic therapy use for ST-segment elevation myocardial infarction and long-term outcomes in China: 2-year results from the China Acute Myocardial Infarction Registry. BMC Cardiovasc Disord. 2023 Feb 22;23(1):103. doi: 10.1186/s12872-023-03105-1. PMID: 36814182; PMCID: PMC9948459.

12. Ministry of Health of People’s Republic of China. China public health statistical yearbook 2021. Beijing: Peking Union Medical College Publishing House; 2021.

13. Ibanez B, James S, Agewall S, Antunes MJ, Bucciarelli-Ducci C, Bueno H, Caforio ALP, Crea F, Goudevenos JA, Halvorsen S, Hindricks G, Kastrati A, Lenzen MJ, Prescott E, Roffi M, Valgimigli M, Varenhorst C, Vranckx P, Widimský P; ESC Scientific Document Group. 2017 ESC Guidelines for the management of acute myocardial infarction in patients presenting with ST-segment elevation: The Task Force for the management of acute myocardial infarction in patients presenting with ST-segment elevation of the European Society of Cardiology (ESC). Eur Heart J. 2018 Jan 7;39(2):119-177. doi: 10.1093/eurheartj/ehx393. PMID: 28886621.

14. Levine GN, Bates ER, Bittl JA, Brindis RG, Fihn SD, Fleisher LA, Granger CB, Lange RA, Mack MJ, Mauri L, Mehran R, Mukherjee D, Newby LK, O’Gara PT, Sabatine MS, Smith PK, Smith SC Jr. 2016 ACC/AHA Guideline Focused Update on Duration of Dual Antiplatelet Therapy in Patients With Coronary Artery Disease: A Report of the American College of Cardiology/American Heart Association Task Force on Clinical Practice Guidelines: An Update of the 2011 ACCF/AHA/SCAI Guideline for Percutaneous Coronary Intervention, 2011 ACCF/AHA Guideline for Coronary Artery Bypass Graft Surgery, 2012 ACC/AHA/ACP/AATS/PCNA/SCAI/STS Guideline for the Diagnosis and Management of Patients With Stable Ischemic Heart Disease, 2013 ACCF/AHA Guideline for the Management of ST-Elevation Myocardial Infarction, 2014 AHA/ACC Guideline for the Management of Patients With Non-ST-Elevation Acute Coronary Syndromes, and 2014 ACC/AHA Guideline on Perioperative Cardiovascular Evaluation and Management of Patients Undergoing Noncardiac Surgery. Circulation. 2016 Sep 6;134(10):e123-55. doi: 10.1161/CIR.0000000000000404. Epub 2016 Mar 29. Erratum in: Circulation. 2016 Sep 6;134(10):e192-4. PMID: 27026020.

15. WU Chao, ZHANG Xiaoyu, YU Mei, et al. Analysis of In-hospital Outcome of Patients With ST-segment Elevation Myocardial Infarction Undergoing Various Thrombolytic Strategies in China, 2021, 36: 1070–1076. DOI: 10.3969/j.issn.1000-3614.2021.11.003.

16. Armstrong PW, Gershlick AH, Goldstein P, Wilcox R, Danays T, Lambert Y, Sulimov V, Rosell Ortiz F, Ostojic M, Welsh RC, Carvalho AC, Nanas J, Arntz HR, Halvorsen S, Huber K, Grajek S, Fresco C, Bluhmki E, Regelin A, Vandenberghe K, Bogaerts K, Van de Werf F; STREAM Investigative Team. Fibrinolysis or primary PCI in ST-segment elevation myocardial infarction. N Engl J Med. 2013 Apr 11;368(15):1379–87. doi: 10.1056/NEJMoa1301092. Epub 2013 Mar 10. PMID: 23473396.

17. Widimsky P, Bilkova D, Penicka M, Novak M, Lanikova M, Porizka V, Groch L, Zelizko M, Budesinsky T, Aschermann M; PRAGUE Study Group Investigators. Long-term outcomes of patients with acute myocardial infarction presenting to hospitals without catheterization laboratory and randomized to immediate thrombolysis or interhospital transport for primary percutaneous coronary intervention. Five years’ follow-up of the PRAGUE-2 Trial. Eur Heart J. 2007 Mar;28(6):679-84. doi: 10.1093/eurheartj/ehl535. Epub 2007 Feb 13. PMID: 17298968.

18. Masoudi FA, Ponirakis A, de Lemos JA, Jollis JG, Kremers M, Messenger JC, Moore JWM, Moussa I, Oetgen WJ, Varosy PD, Vincent RN, Wei J, Curtis JP, Roe MT, Spertus JA. Trends in U.S. Cardiovascular Care: 2016 Report From 4 ACC National Cardiovascular Data Registries. J Am Coll Cardiol. 2017 Mar 21;69(11):1427-1450. doi: 10.1016/j.jacc.2016.12.005. Epub 2016 Dec 23. PMID: 28025065.

19. Zhao L, Zhao Z, Chen X, Li J, Liu J, Li G; Group of Prourokinase Phase IV Clinical Trials Investigators. Safety and efficacy of prourokinase injection in patients with ST-elevation myocardial infarction: phase IV clinical trials of the prourokinase phase study. Heart Vessels. 2018 May;33(5):507–512. doi: 10.1007/s00380-017-1097-x. Epub 2017 Dec 5. PMID: 29209778.

20. Liu Y, Yang Y, Li Y, Peng X. Comparison of Efficacy and Safety of Recombinant Human Prourokinase and Alteplase in the Treatment of STEMI and Analysis of Influencing Factors of Efficacy. Evid Based Complement Alternat Med. 2021 Sep 6;2021:6702965. doi: 10.1155/2021/6702965. PMID: 34531919; PMCID: PMC8440075.

21. Virmani R, Kolodgie FD, Burke AP, Farb A, Schwartz SM. Lessons from sudden coronary death: a comprehensive morphological classification scheme for atherosclerotic lesions. Arterioscler Thromb Vasc Biol 2000;20:1262–75.

22. Jia H, Abtahian F, Aguirre AD, et al. In vivo diagnosis of plaque erosion and calcified nodule in patients with acute coronary syndrome by intravascular optical coherence tomography. J Am Coll Cardiol 2013;62: 1748–58.

23. Fernandez-Ortiz A, Badimon JJ, Falk E, et al. Characterization of the relative thrombogenicity of atherosclerotic plaque components: implications for consequences of plaque rupture. J Am Coll Cardiol 1994;23:1562–9.

24. Hu S, Yonetsu T, Jia H, Karanasos A, Aguirre AD, Tian J, Abtahian F, Vergallo R, Soeda T, Lee H, McNulty I, Kato K, Yu B, Mizuno K, Toutouzas K, Stefanadis C, Jang IK. Residual thrombus pattern in patients with ST-segment elevation myocardial infarction caused by plaque erosion versus plaque rupture after successful fibrinolysis: an optical coherence tomography study. J Am Coll Cardiol. 2014 Apr 8;63(13):1336–1338. doi: 10.1016/j.jacc.2013.11.025. Epub 2013 Dec 18. PMID: 24361315.

25. Sinnaeve PR, Armstrong PW, Gershlick AH, Goldstein P, Wilcox R, Lambert Y, Danays T, Soulat L, Halvorsen S, Ortiz FR, Vandenberghe K, Regelin A, Bluhmki E, Bogaerts K, Van de Werf F; STREAM investigators. ST-segment-elevation myocardial infarction patients randomized to a pharmaco-invasive strategy or primary percutaneous coronary intervention: Strategic Reperfusion Early After Myocardial Infarction (STREAM) 1-year mortality follow-up. Circulation. 2014 Sep 30;130(14):1139-45. doi: 10.1161/CIRCULATIONAHA.114.009570. Epub 2014 Aug PMID: 25161043.

26. Pu J, Ding S, Ge H, Han Y, Guo J, Lin R, Su X, Zhang H, Chen L, He B; EARLY-MYO Investigators. Efficacy and Safety of a Pharmaco-Invasive Strategy With Half-Dose Alteplase Versus Primary Angioplasty in ST-Segment-Elevation Myocardial Infarction: EARLY-MYO Trial (Early Routine Catheterization After Alteplase Fibrinolysis Versus Primary PCI in Acute ST-Segment-Elevation Myocardial Infarction). Circulation. 2017 Oct 17;136(16):1462–1473. doi: 10.1161/CIRCULATIONAHA.117.030582. Epub 2017 Aug 27. Erratum in: Circulation. 2018 Feb 13;137(7):e29. PMID: 28844990.

27. Armstrong P. Pharmaco-invasive reperfusion with half-dose tenecteplase or primary PCI in older patients with STEMI. Presented at: ACC/WCC 2023. March 5, 2023. New Orleans, LA.

28. Jortveit J, Pripp AH, Halvorsen S. Outcomes after delayed primary percutaneous coronary intervention vs. pharmaco-invasive strategy in ST-segment elevation myocardial infarction in Norway. Eur Heart J Cardiovasc Pharmacother. 2022 Aug 11;8(5):442–451. doi: 10.1093/ehjcvp/pvab041. PMID: 34038535; PMCID: PMC9366642.

29. Siddiqi TJ, Usman MS, Khan MS, Sreenivasan J, Kassas I, Riaz H, Raza S, Deo SV, Sharif H, Kalra A, Yadav N. Meta-Analysis Comparing Primary Percutaneous Coronary Intervention Versus Pharmacoinvasive Therapy in Transfer Patients with ST-Elevation Myocardial Infarction. Am J Cardiol. 2018 Aug 15;122(4):542–547. doi: 10.1016/j.amjcard.2018.04.057. Epub 2018 Jun 20. PMID: 30205885.

30. Jamal J, Idris H, Faour A, Yang W, McLean A, Burgess S, Shugman I, Wales K, O’Loughlin A, Leung D, Mussap CJ, Juergens CP, Lo S, French JK. Late outcomes of ST-elevation myocardial infarction treated by pharmaco-invasive or primary percutaneous coronary intervention. Eur Heart J. 2023 Feb 7;44(6):516–528. doi: 10.1093/eurheartj/ehac661. PMID: 36459120.

31. Liu J, Fu XH, Xue L, Wu WL, Gu XS, Li SQ. Equilibrium radionuclide angiography for evaluating the effect of facilitated percutaneous coronary intervention on ventricular synchrony in patients with acute myocardial infarction. Circ J. 2012;76(4):928–35. doi: 10.1253/circj.cj-11-1329. Epub 2012 Feb 7. PMID: 22313803.

